# The prevalence of mental ill-health in women during pregnancy and after childbirth during the Covid-19 pandemic: a Systematic review and Meta-analysis

**DOI:** 10.1101/2022.06.13.22276327

**Authors:** Gayathri Delanerolle, Mary McCauley, Martin Hirsch, Yutian Zheng, Xu Cong, Heitor Cavalini, Ashish Shetty, Shanaya Rathod, Jian Qing Shi, Dharani K Hapangama, Peter Phiri

## Abstract

**Background:** Severe Acute Respiratory Syndrome Coronavirus (SARS-CoV) is a respiratory disease causing coronavirus. SARS-CoV has caused the Middle East Respiratory Syndrome (MERS), SARS-CoV in Hong King and SARS-CoV-2 (COVID-19). COVID-19, to date, have had the highest mortality and morbidity globally, thus reaching the pandemic status. In comparison to research conducted to explore the impact of pandemics on the general wellbeing, there appears to be a paucity on its association with women’s mental health. Many pregnant women have reported that the pandemic negatively impacted their mental health.

**Aim:** This study aimed is to explore the prevalence of the impact of the COVID-19, MERS and SARS pandemics on the mental health of pregnant women.

**Method:** A study protocol was developed and published in PROSPERO (CRD42021235356) to explore a number of key objectives. For the purpose of this study PubMed, Science direct, Ovid PsycINFO and EMBASE databases were searched from December 2000 – July 2021. The search results were screened, first by title, and then by abstract. A meta-analysis was conducted to report the findings.

**Results:** There were no studies reporting the mental health impact due to MERS and SARS. We systematically identified 316 studies that reported on the mental health of women that were pregnant and soon after birth. The meta-analysis indicated 24.9% (21.37%-29.02%) of pregnant women reported symptoms of depression, 32.8% (29.05% to 37.21%) anxiety, 29.44% (18.21% - 47.61%) stress, 27.93% (9.05%-86.15 %) PTSD, and 24.38% (11.89%-49.96%) sleep disorders during the COVID-19 pandemic. Furthermore, the I^2^ test showed a high heterogeneity value.

**Conclusion:** The importance of managing the mental health during pregnancy and after-delivery improves the quality of life and wellbeing of mothers. Developing an evidence based mental health framework as part of pandemic preparedness to help pregnant women would improve the quality of care received during challenging times.

## Background

Since December 2019, the coronavirus disease 2019 (COVID-19) pandemic caused by severe acute respiratory syndrome coronavirus 2 (SARS-Cov-2) has spread around the world unprecedentedly, overwhelming healthcare systems around the world. On March 11, 2020, the World Health Organization (WHO) declared COVID-19 a global pandemic. This led to a rippling impact of the virus on healthcare systems and patients who needed to access care for both physical and mental health and wellbeing [1]. There was concern that the acute intensive care services would not be able to cope with the growing volume of affected individuals requiring ventilatory support. To reduce viral transmission and relieve pressure on healthcare systems, many countries, including the United Kingdom (UK), entered lockdown.

People were ordered by law to stay at home. In many hospitals, staff were redeployed and departments were adapted or converted to COVID-19 services. However, women who were pregnant and needed to give birth were identified as a vulnerable group and the ability to provide good quality maternity care during the Covid-19 pandemic was prioritised.

It is well documented that public health emergencies not only have a huge impact on the physical health of a population but also results in an increase in mental ill-health including: conditions such as depression; post-traumatic stress disorder (PTSD); substance use disorder; behavioural disorders; noncompliance with public health directives, domestic violence; and child abuse[5]. These can arise from triggers directly related to the infection, for example, the neuroinvasive potential of SARS-CoV-2 may affect brain function and mental health. The treatment for COVID-19 may also have adverse effects on mental health and indirectly may contribute to anxiety. In addition, the imposition of unfamiliar and undesired public health measures including social isolation strongly correlates with the likelihood of clinically significant depression or anxiety [5,6]. These findings were echoed in an evaluation of severe acute respiratory syndrome (SARS) epidemic with increases in PTSD, stress, and psychological distress in both patients and clinicians. Affected individuals and communities were motivated to comply with quarantine to reduce the risk of infecting others and to protect their community’s health. However emotional distress tempted some to consider violating the recommended public health measures[6].

One such vulnerable group is women during their pregnancy and after childbirth. Maternal mental ill-health has been an international public health concern for many years[1] with millions of women experiencing mental ill-health during pregnancy and after childbirth[1, 2]. Common mental disorders (depression, anxiety) rank third in the list of the burden of disease globally and maternal mental ill-health affects up to 10% of women during pregnancy and 13% of women after childbirth[6, 7]. It is well documented that compromised maternal mental ill-health is associated with adverse short and long-term consequences for the mother and the baby[12, 13]. However, limited data exists on the prevalence of mental ill-health in women who were pregnant and gave birth during the COVID-19 pandemic. This systematic review and meta-analysis therefore assessed the prevalence of mental ill-health in women during pregnancy and after childbirth during the Covid-19 pandemic. We then compared our findings in relation to other global pandemics including severe acute respiratory syndrome (SARS) and Middle Eastern Respiratory Syndrome (MERS).

## Methods

A systematic methodology was developed along with a relevant protocol that was peer reviewed and published in PROSPERO (CRD42021235356). The developed method focuses on the prevalence of mental ill-health in women during pregnancy and after childbirth during the Covid-19 pandemic.

### Search criteria

The search criteria was developed based upon the research question using PubMed, Science direct, Ovid PsycINFO and EMBASE databases: PubMed, Science direct and EMBASE. We developed a wide search criterion to ensure the inclusion of any pregnant women with existing gynaecological conditions. The MeSH terms used include (COVID) OR (SARS-CoV-2) AND (SARS) AND (MERS) AND ((mental health) OR (depression) OR (anxiety) OR (PTSD) OR (psychosis) OR (unipolar) OR (bipolar)) AND ((PCOS) OR (fibroid) OR (endometriosis) OR (pre-eclampsia) OR (still birth) OR (GDM) OR (preterm birth) OR (women’s health) OR (pregnant women) OR (pregnancy)).

### Screening eligibility criteria

All studies published in English were included from 20^th^ December 2019 to 31^st^ July 2021. Screening and data extraction were performed by two authors independently. Initially, titles and abstracts were reviewed to determine the relevance. A PRISMA diagram was completed based on the eligibility steps completed.

### Data extraction

Full texts of the included papers were reviewed carefully to extract data including time and locations of the study, participants and sample size, mean age, gestation, days since childbirth, prevalence of mental symptoms, data collection tools used, and cut-offs scores applied. Any disagreement was discussed and resolved by consensus between two authors. For studies with both COVID-19 cohort and non-COVID-19 cohort, we only used data of the COVID-19 cohort and the p-value comparing them. Studies from SARS and MERS were also reviewed in full to ensure the eligibility criteria was met. For studies reporting mean (SD) or median (IQR) of the scales measuring mental symptoms instead of prevalence rates were included and a simulation method assuming normal distribution was applied to generate the corresponding prevalence rates.

### Risk of bias assessment

A risk of bias assessment was completed with a RoB table.

### Data analysis

Random effects model with restricted maximum-likelihood estimation method was applied for meta-analysis and I-square statistic was used to evaluate heterogeneity across studies. The pooled prevalence rates of anxiety, depression, PTSD, stress and sleep disorder with 95% confidence interval (CI) were computed. Subgroup analysis was conducted in terms of trimester. Sensitivity analysis was performed to assess the robustness of the results. Potential publication bias was assessed with funnel plot and Egger’s test. Analyses were conducted with the R studio (version 1.4.17.17) and STATA 16.1.

## Results

Our initial search identified a total of 1603 papers and 523 studies were excluded after screening by titles and abstracts. After full-text evaluation, 217 and 99 studies were included in the systematic review and meta-analysis, respectively. The PRISMA (Preferred Reporting Items for Systematic Reviews and Meta-analyses) flowchart was illustrated in **Figure 1**.

**Figure 1.**
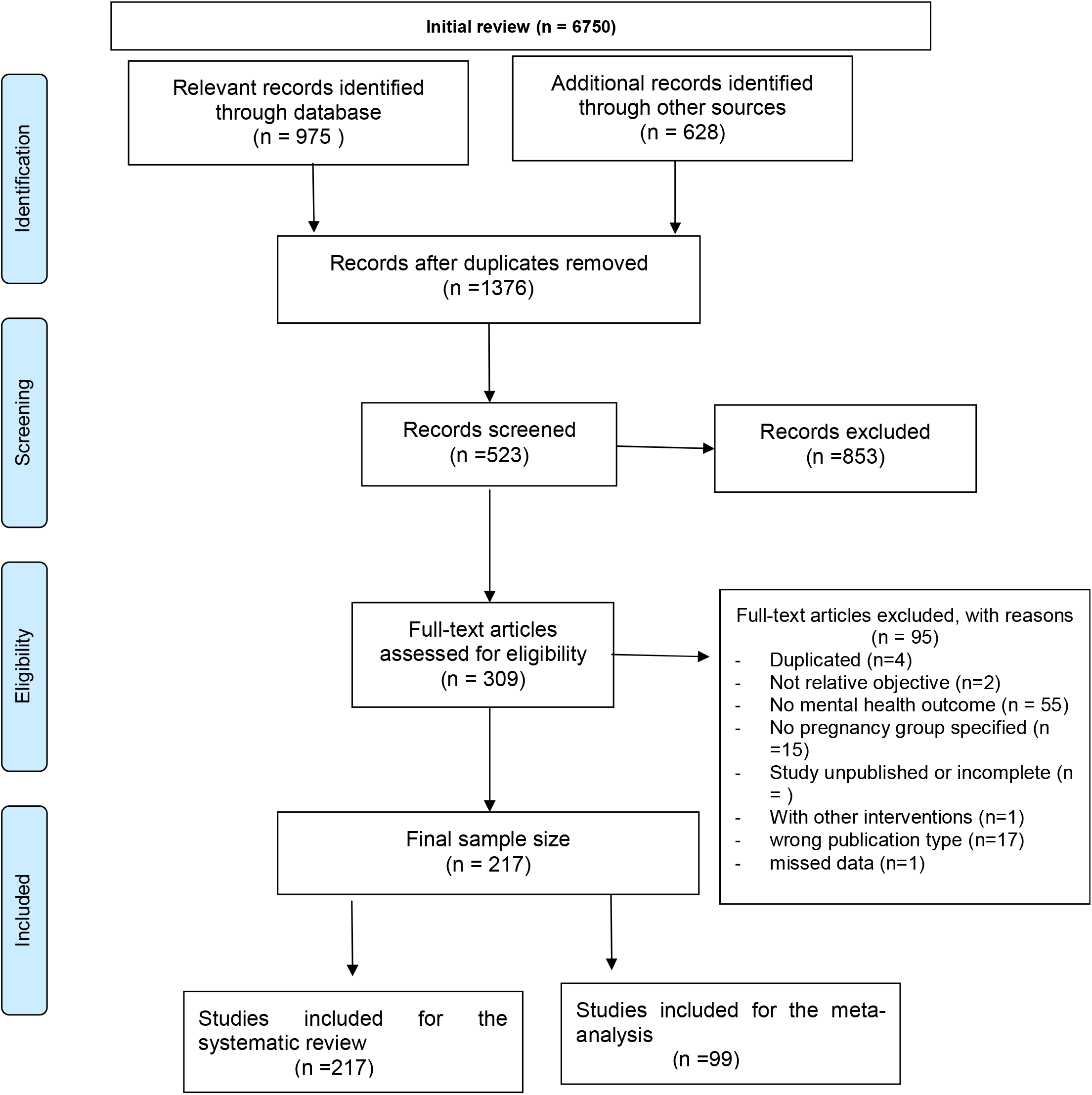
PRISMA Flow Diagram.

### Inclusion and exclusion criteria

All COVID-19, SARS and MERS studies that evaluated the mental health of pregnant women that may or may not have gynaecological conditions that were reported in English between December 2000 – July 2021 were included. All other studies were excluded from this analysis.

### Characteristics of studies

A total of 217 COVID-19 studies were included and 99 studies were meta-analysed. These studies were reported from various parts of the world, as indicated in the characteristics **Table 1**. We did not identify SARS and MERS studies that were suitably aligned to the eligibility criteria of our study.

**Table 1.**
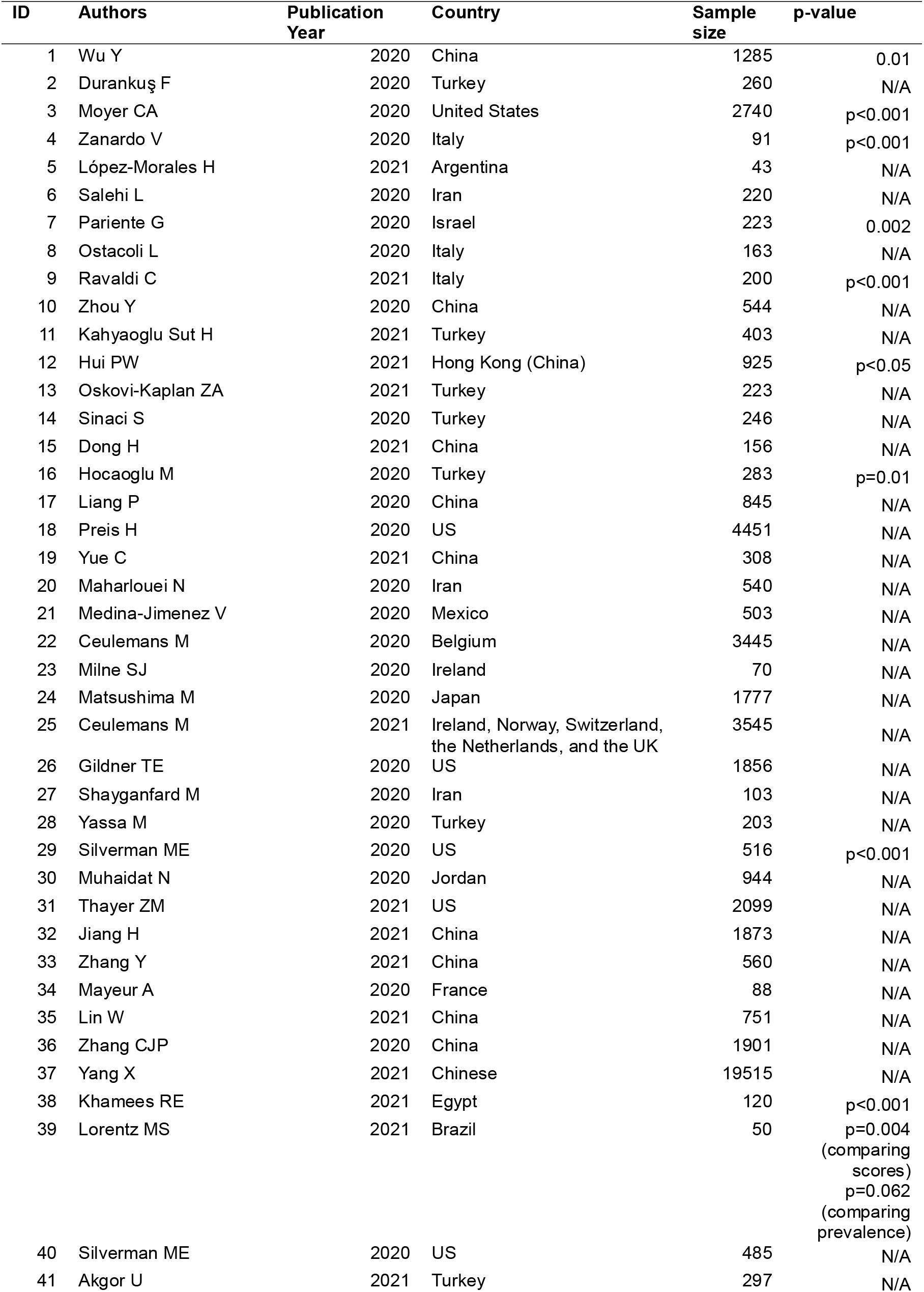
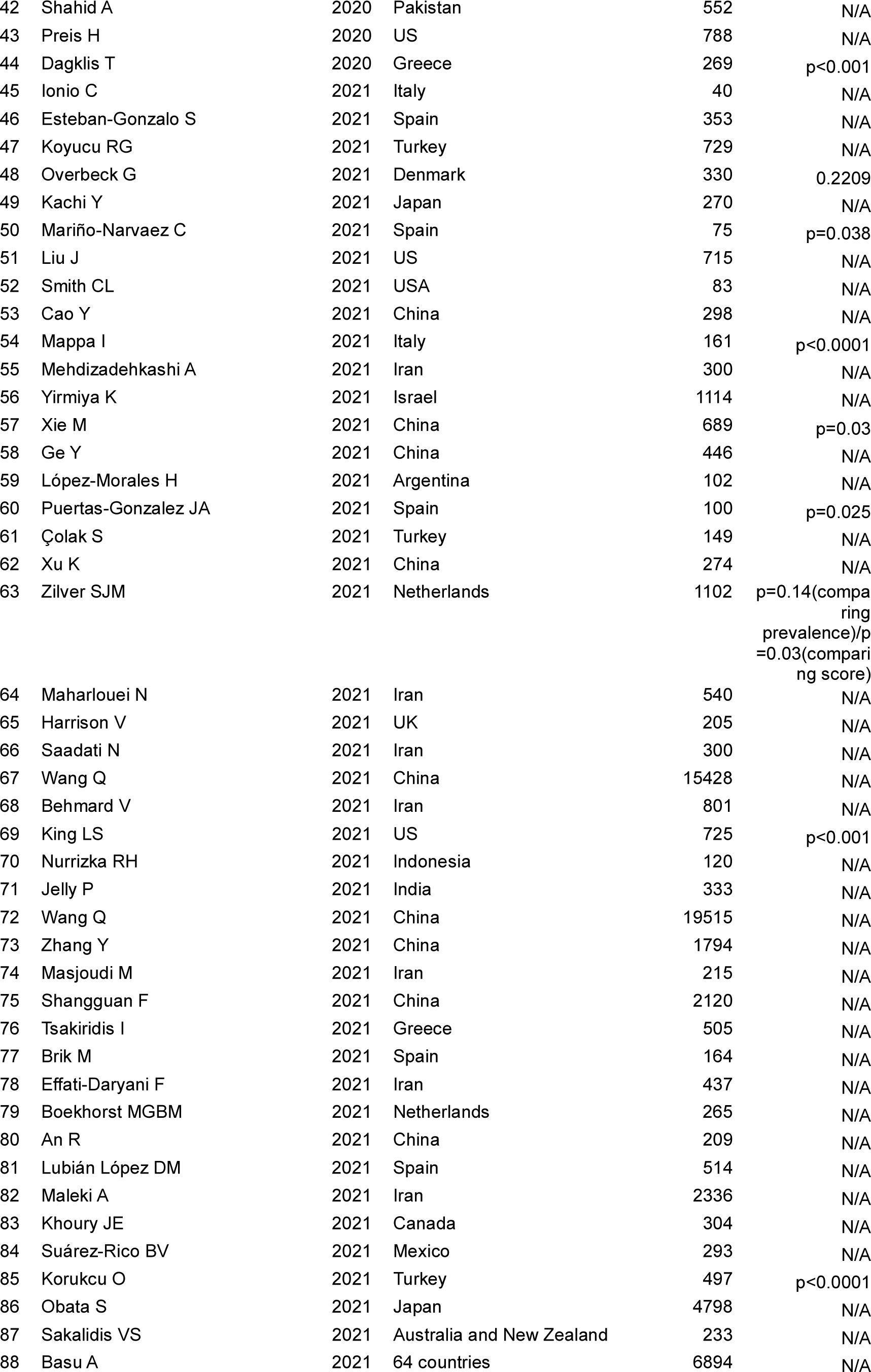
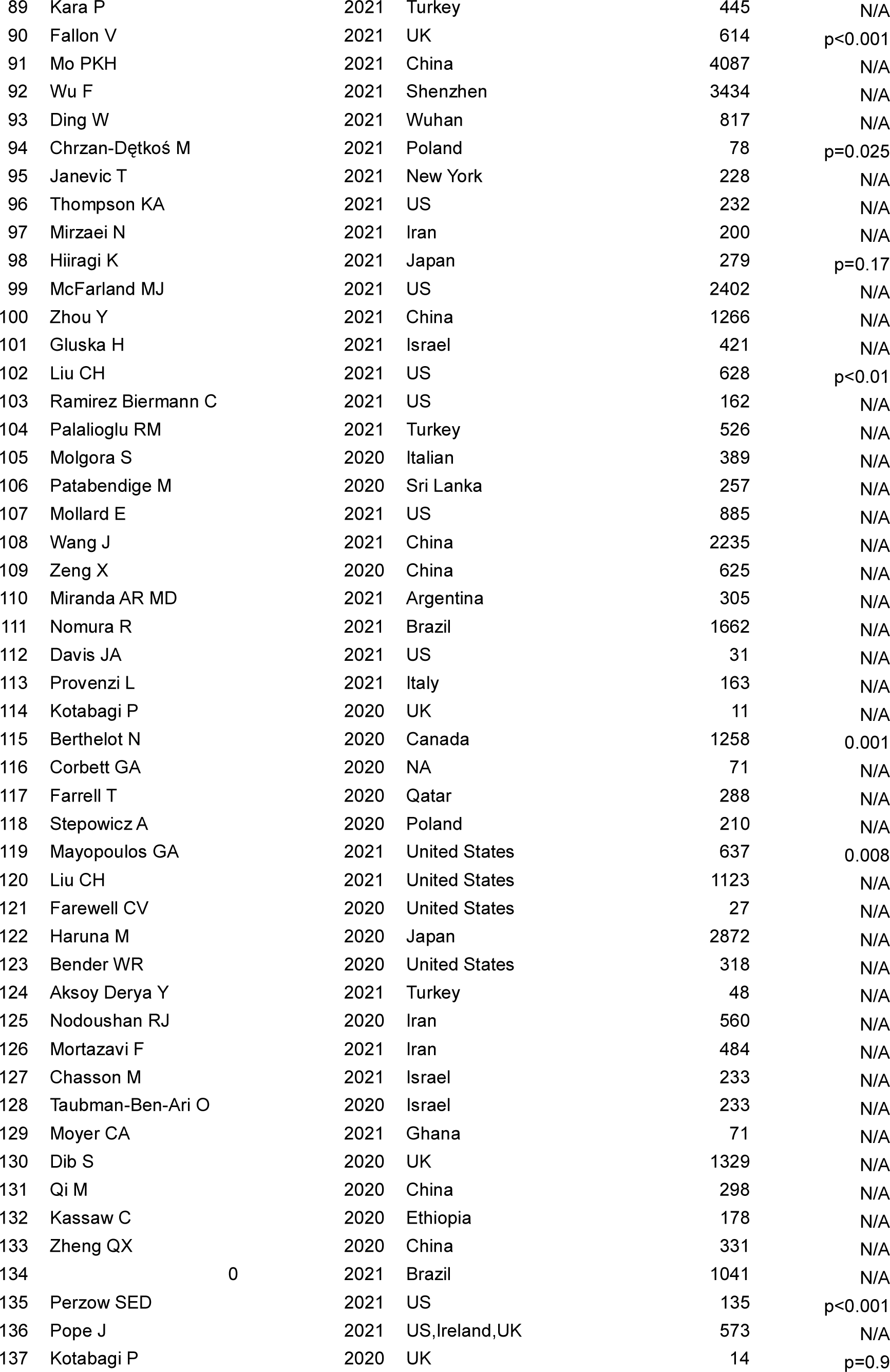
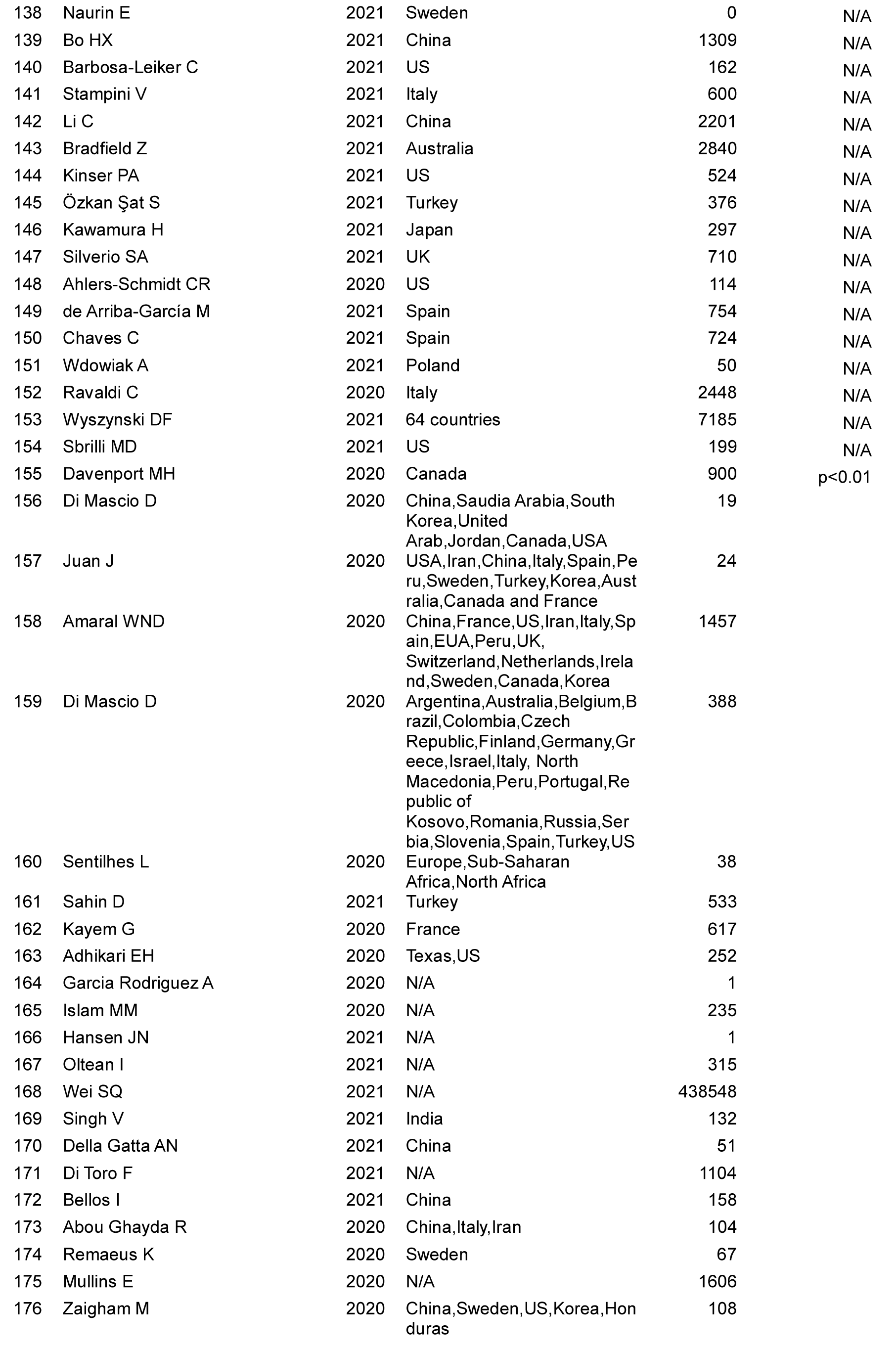
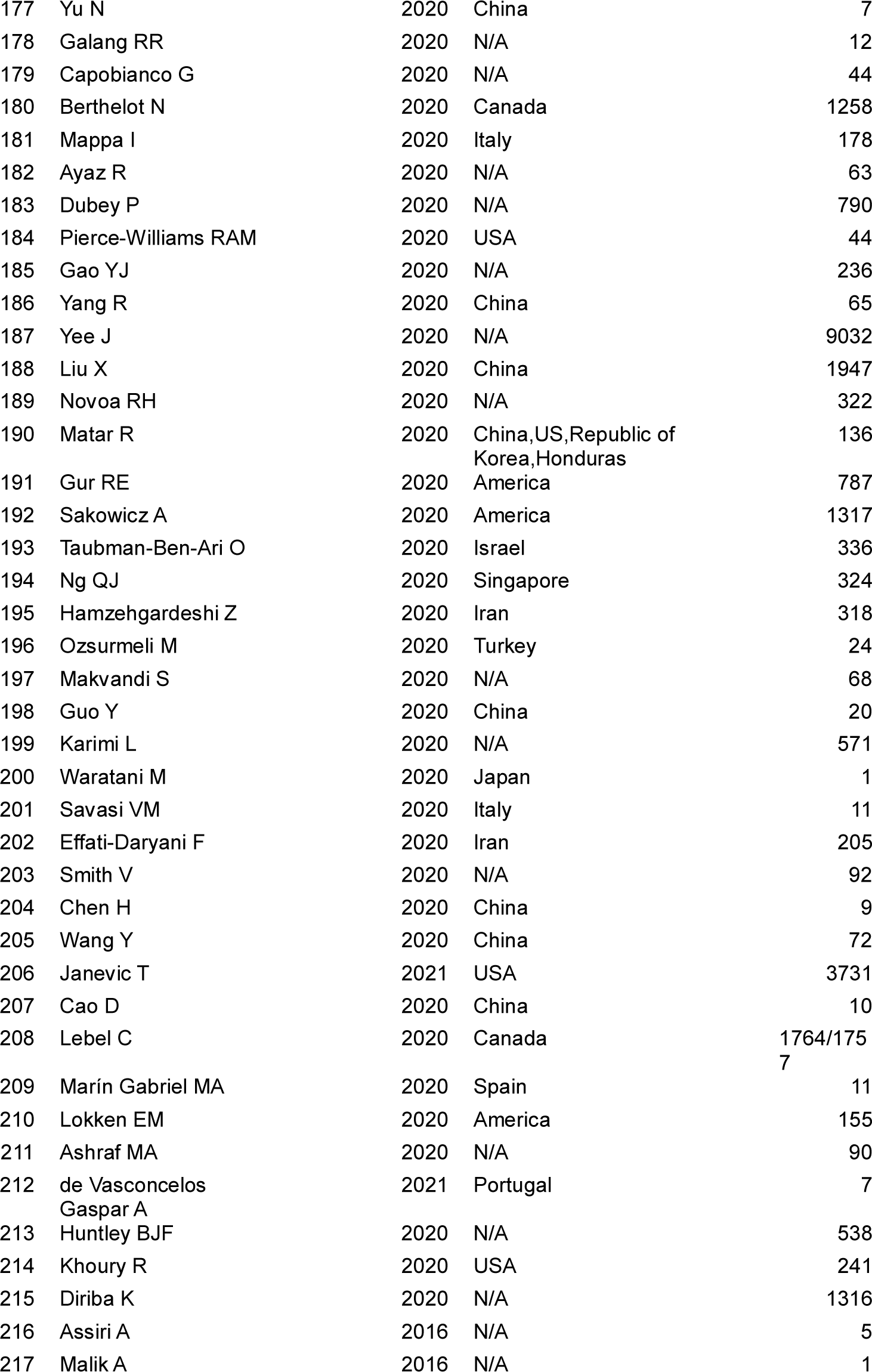
217 studies in systematic review and meta-analysis.

#### Study design, source of data, data collection method and sample size

All 217 studies used different study designs; 107 cross-sectional, 7 cohort and 7 case controlled. A total of 23 qualitative studies used self-reported methods of data collection. All studies reported a variety of mental health symptoms. Real-world data from hospital admissions were used in 5 studies whilst 2 extracted data from patient medical records. The 217 study pool comprised of a sample of 638,889 pregnant women whilst 6898 were within 90 days of delivery. The sample sizes used within the studies varied considerably; 129 comprised of approximately 500, 40 with 500–999, 18 with 1000–1999 and 24 ≥2000 women.

#### Stages of pregnancy assessed

A total of 99 studies reported pregnant women during their first, second and third trimester.

#### Site of data collection

Many studies reported that data collection took place during routine antenatal or postnatal visits in outpatient departments, tertiary/provincial hospitals, secondary level or district hospitals and primary healthcare facility level.

Of the 217 systematically included studies, 64 reported data on depression, 82 on anxiety, 20 on stress, 7 on PTSD, and 8 on sleep disorder. Detailed characteristics of the systematically included studies and those meta-analysed are listed in **Table 1 and 2 (table 2 supplementary material)**.

**Table 2.**
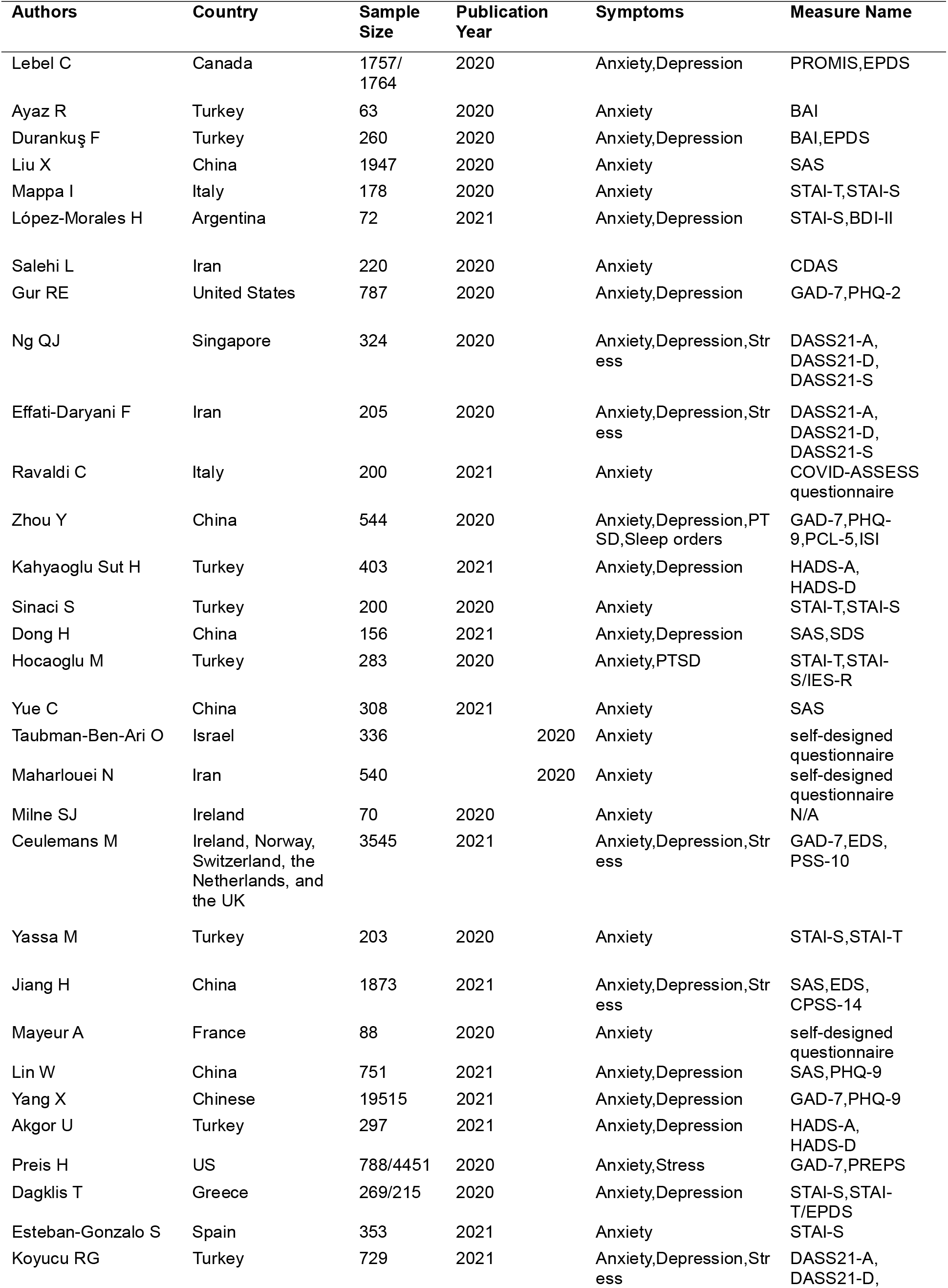
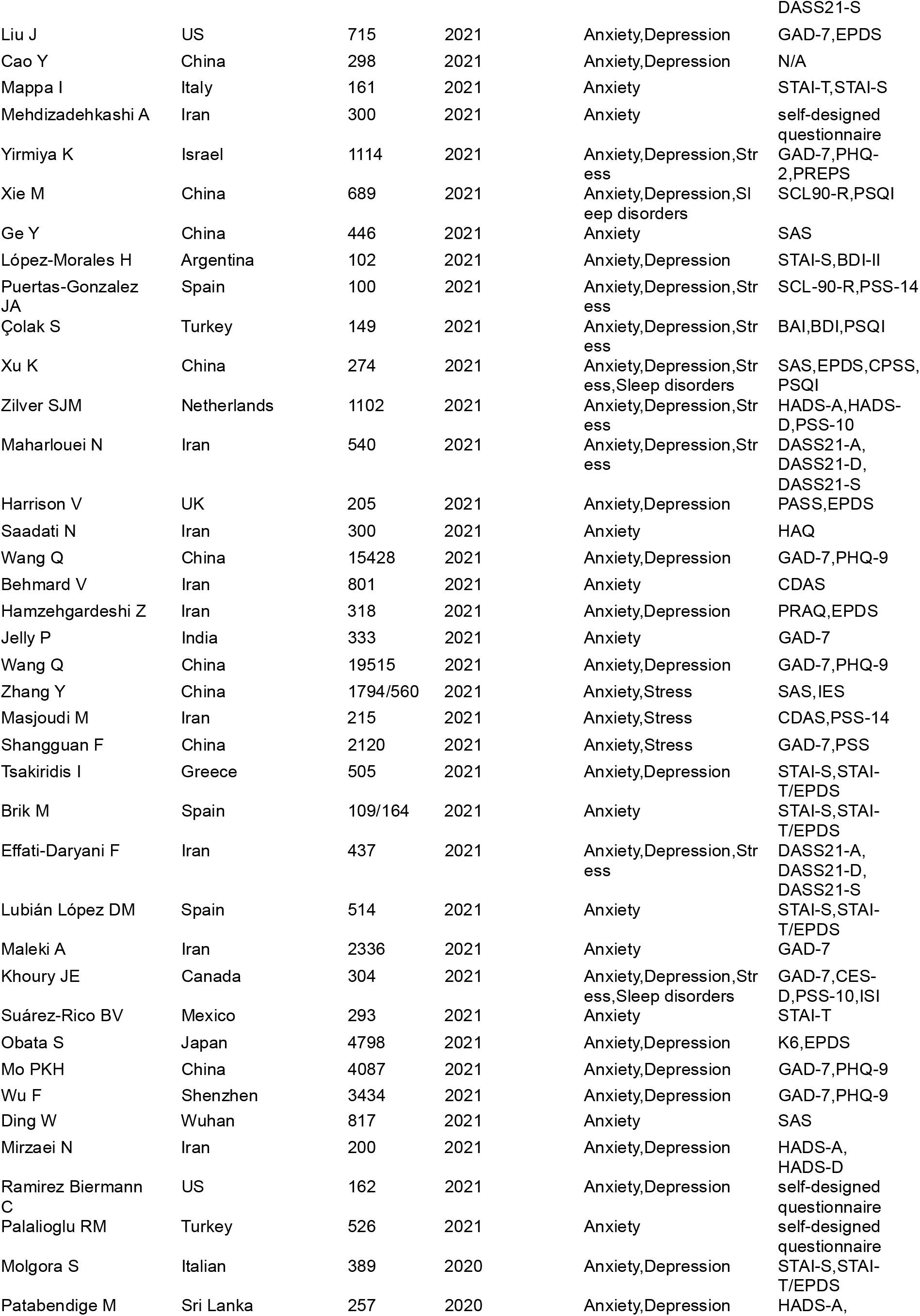
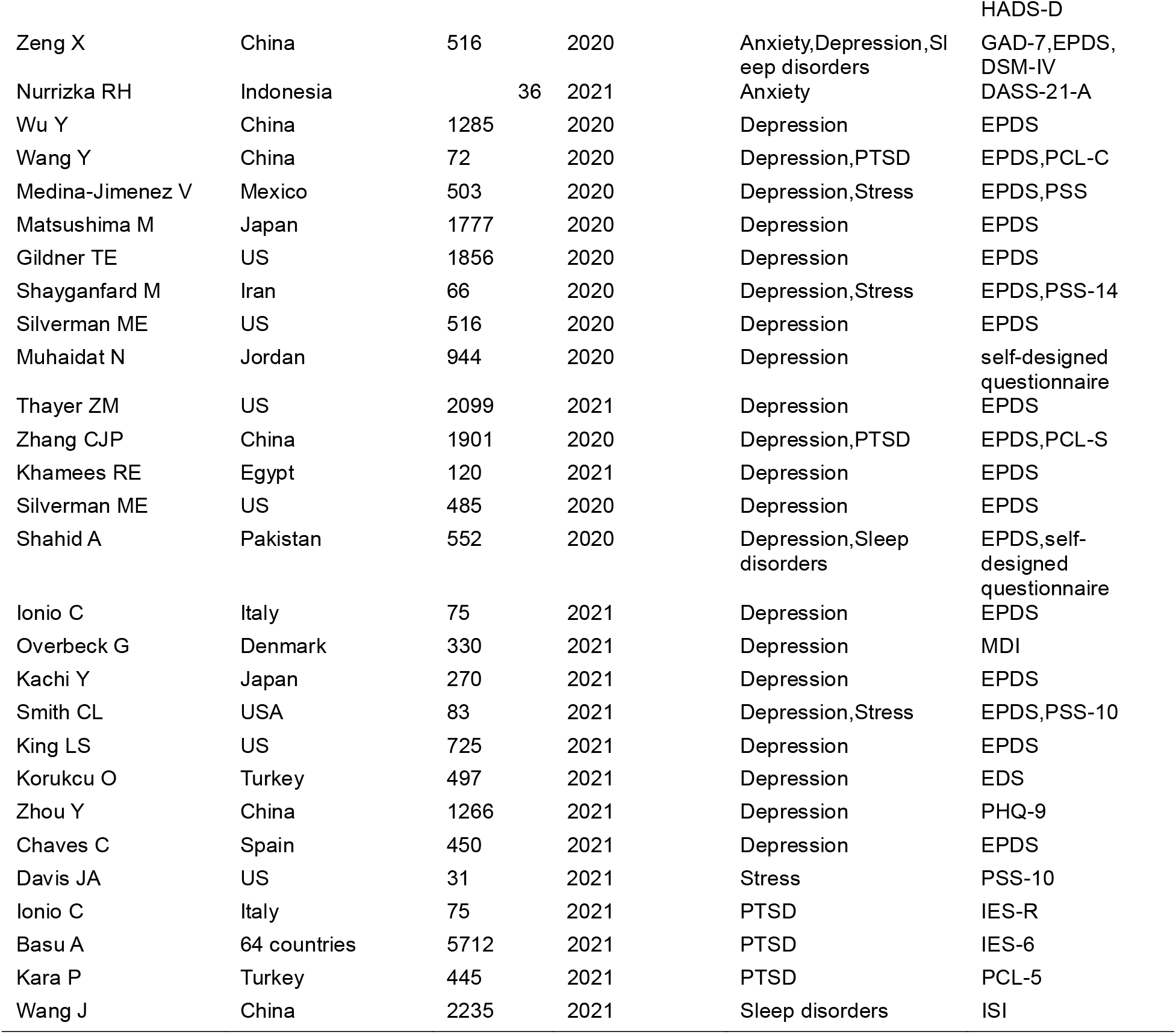
(Supplementary material) 99 studies selected for meta-analysis of depression, anxiety, stress, PTSD and sleep disorders.

#### Meta-analysis

Edinburgh Postnatal Depression Scale (EPDS), the Patient Health Questionnaire 9-item (PHQ-9), the depression subscale of the Hospital Anxiety and Depression Scale (HADS-D) were the commonly used data collection tools to assess symptoms of depression in women during pregnancy and after childbirth. The pooled prevalence of depression was 24.91% with a 95% CI of 21.37%-29.02% (**Figure 2)**.

**Figure 2.**
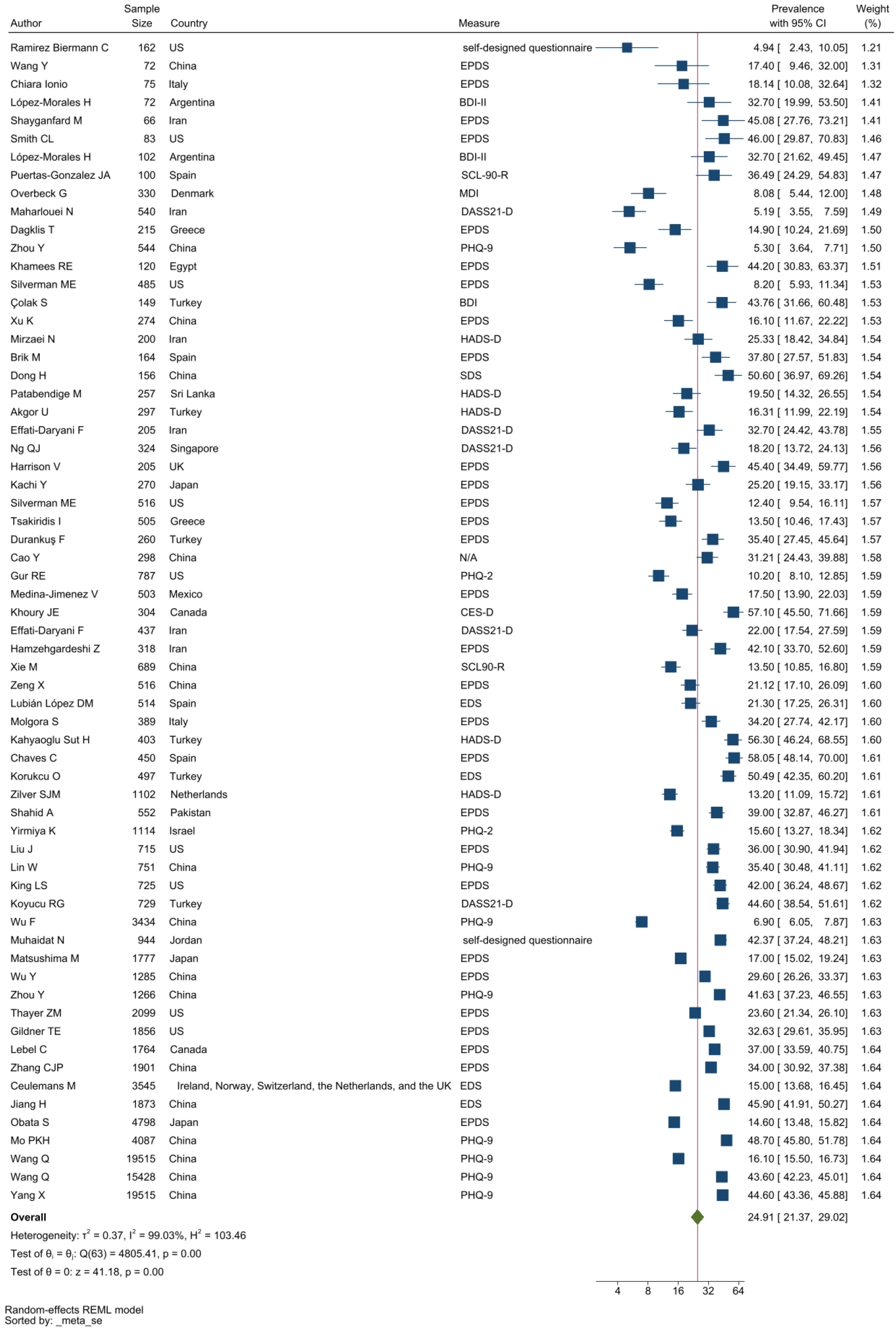
Forest plot of depression.

Anxiety symptoms were commonly measured by the State-Trait Anxiety Inventory (STAI, with two subscales STAI-T and STAI-S), the General Anxiety Disorder 7-item (GAD-7) and Self-rating Anxiety Scale (SAS). Anxiety prevalence was 32.88% with a 95% CI of 29.05% to 37.21% (**Figure 3)**.

**Figure 3.**
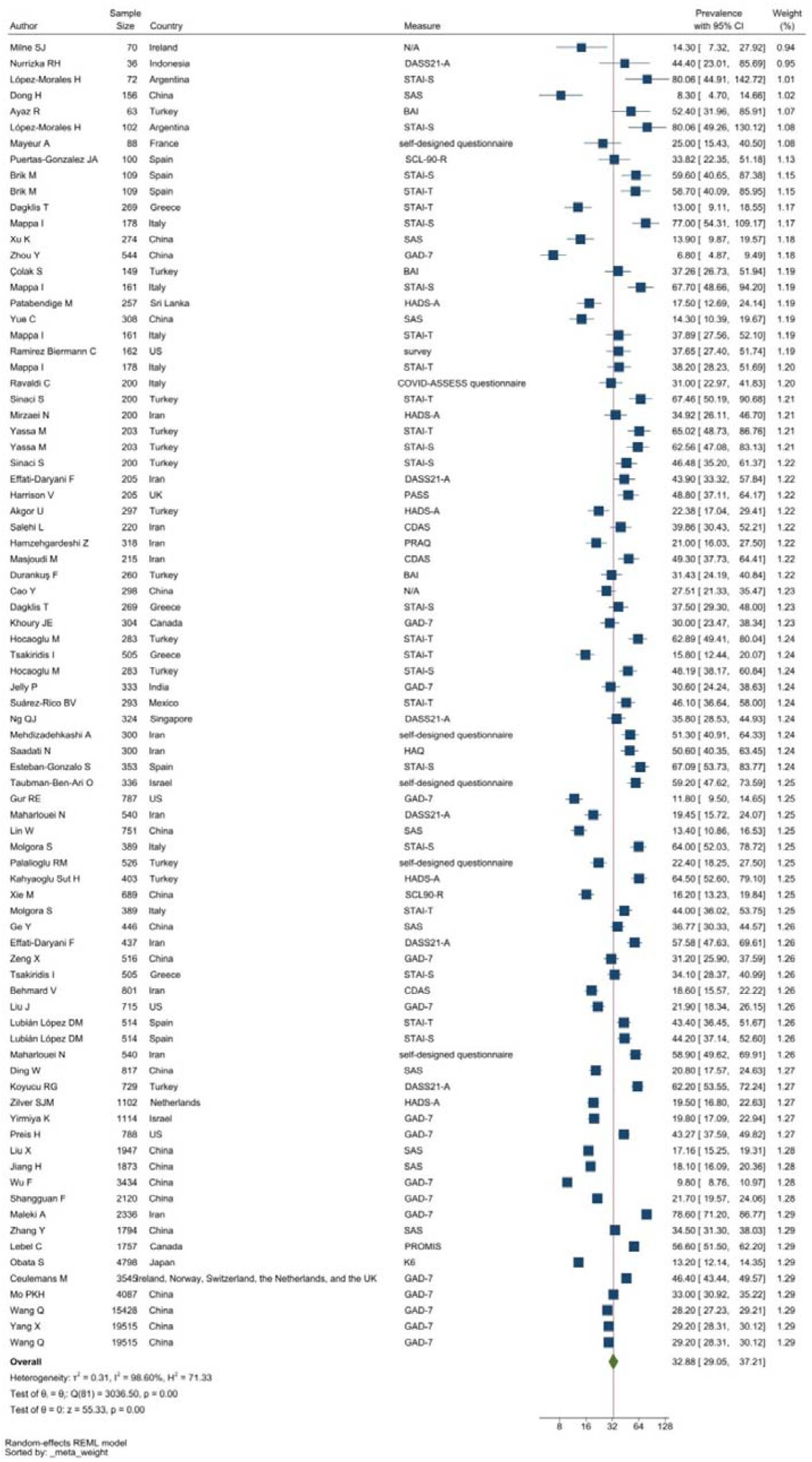
Forest plot of anxiety.

Tools like the Perceived Stress Scale (PSS, with 10-item and 14-item versions), the stress subscale of the 21-item Depression Anxiety and Stress Scale (DASS21-S) were frequently used to evaluate stress symptoms. The pooled prevalence of stress among perinatal women was 29.44% (95% CI: 18.21% - 47.61%) as demonstrated in **Figure 4**.

**Figure 4.**
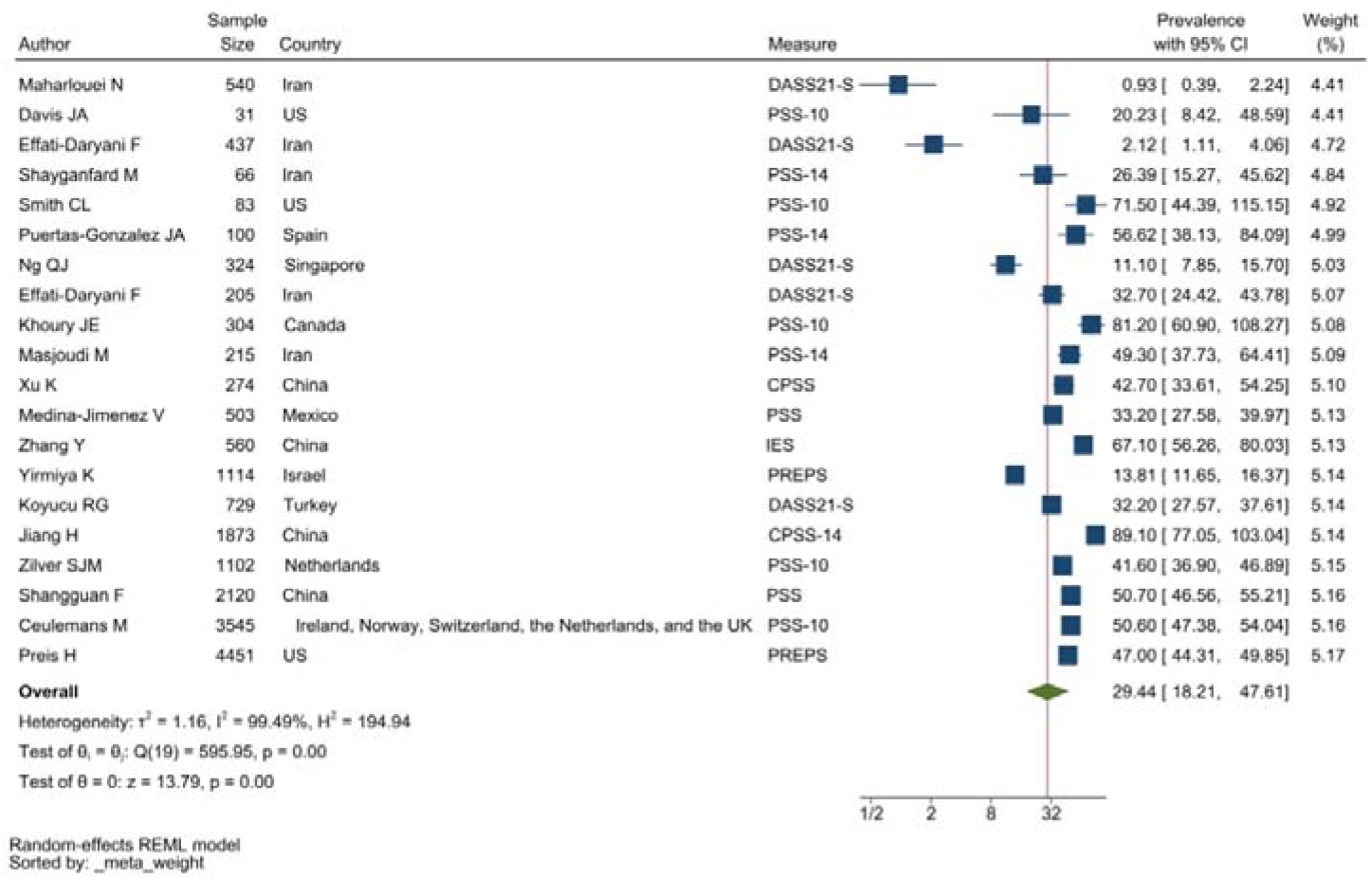
Forest plot of stress.

PTSD symptoms were typically measured by the DSM-V Post-Traumatic Stress Disorder Checklist (PCL-5) and the Impact of Events Scale (IES). The studies reporting PTSD symptoms were heterogeneous (**Figure 5**) resulting in a pooled prevalence of 27.93% with a 95%CI of 9.05%-86.15%.

**Figure 5.**
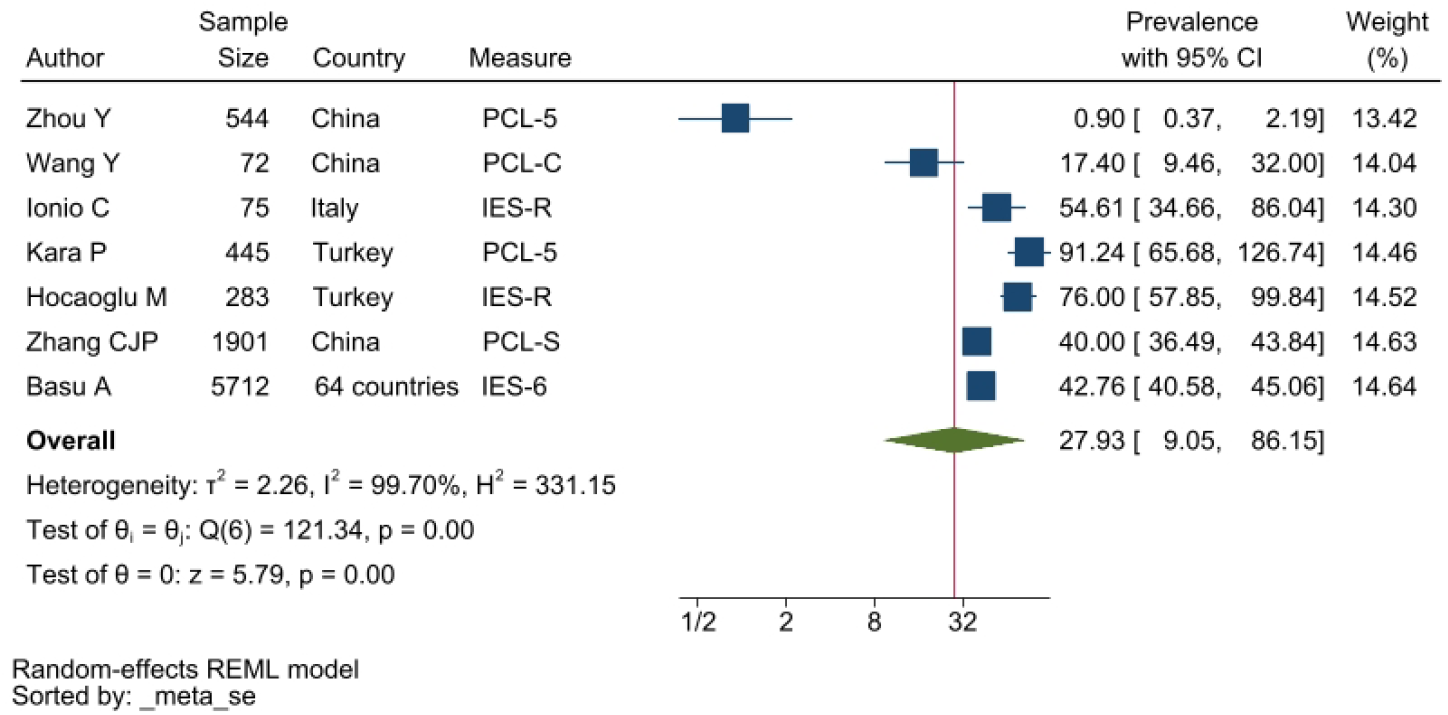
Forest plot of PTSD.

The Insomnia Severity Index (ISI) and the Pittsburgh Sleep Quality Index (PSQI) were to assess and report symptoms associated with sleep disorders. The pooled prevalence was 24.38% with a 95% CI of 11.89%-49.96% (**Figure 6 supplementary material)**.

**Figure 6.**
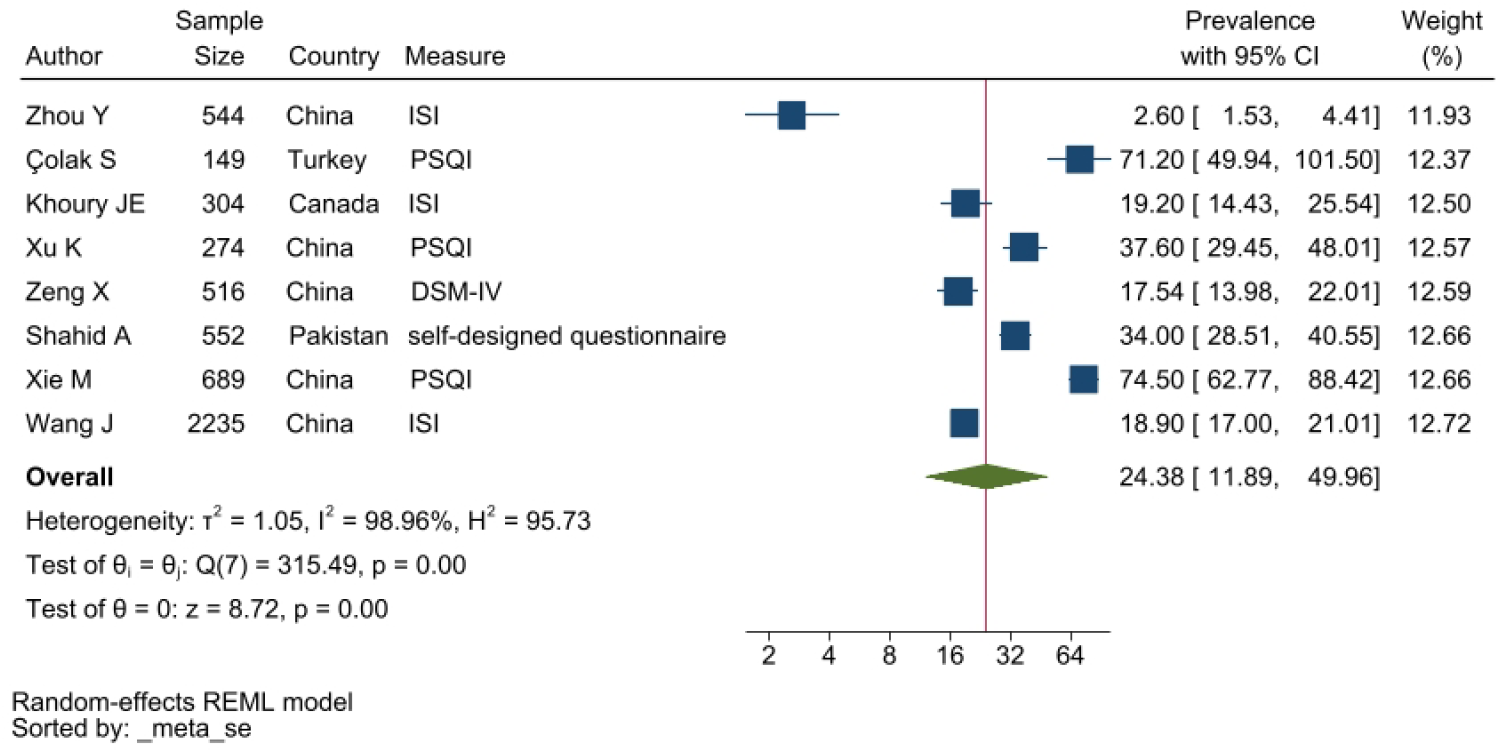
(supplementary material) Forest plot of sleep disorders.

The I^2^ evaluated for depression, anxiety, PTSD, stress and sleep issues were over 98%, which demonstrates a high heterogeneity among the studies. Therefore, a subgroup analysis was conducted to further evaluate the heterogeneity.

#### Subgroup analysis

Women were assessed at different stages of their pregnancy. To determine the rates of depression, anxiety, PTSD, stress and sleep problems, the dataset was categorised based on the trimesters;1^st^ trimester (<12 weeks), 2^nd^ trimester (13-27 weeks), 3^rd^ trimester (28-41 weeks)] and the immediate post-partum period (immediately after childbirth and up to six weeks) for studies that reported follow-up details.

The heterogeneity of depression is lower in comparison to anxiety, PTSD, stress and sleep problems. Heterogeneity within the 1^st^ trimester was 89.47%. I^2^ of the anxiety group during the 1^st^ trimester and 2^nd^ trimester were 88.91% and 92.35%, respectively. These appear to similar to the I^2^ values of depression. I^2^ for stress associated with the 2^nd^ and 3^rd^ trimesters were 78.57% and 64.65%, respectively, indicating mild heterogeneity. Intuitively, Maharlouei and colleagues study reported a small prevalence, thus could be an influencing factor for the heterogeneity reported. I^2^ for PTSD across three trimesters were 24.67%, 89.47% and 81.62%, respectively. I^2^ was 0% during the 1^st^ trimester within the groups of participants reporting sleep disturbance. 1^st^ trimester group showed relatively low heterogeneity across mental health symptoms, thus strictly stipulating the gestational weeks of the included pregnancy helped reduce the heterogeneity.

#### Publication bias and sensitivity analysis

Publication bias and a sensitivity analysis was conducted to assess the reliability of the data as some studies had large standard errors that would produce undesirable effects. Copas selection model was used to select studies for the sensitivity analysis. The p-values of residual selection bias were evaluated as demonstrated in **Figure 17-21**. Studies with a p-value of >0.1 indicated that the residual selection had minimal bias and, the selected studies can be represented. The proportions identified were 67.84%, 100% and 59.49% for depression, anxiety and sleep disorders, respectively. Studies reporting stress and PTSD, the copas selection model could not provide a decision indicating the previous conclusions of high heterogeneity is accurate.

**Figure 7.**
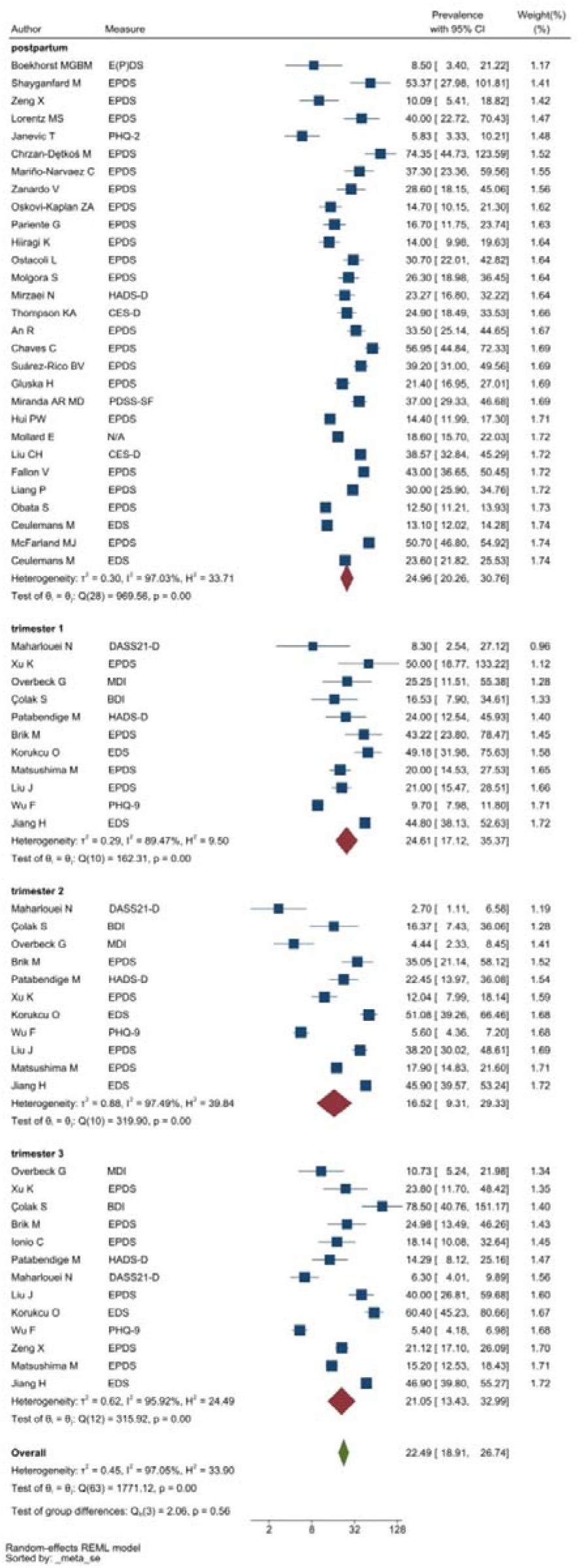
Subgroup analysis of depression (supplementary material)

**Figure 8.**
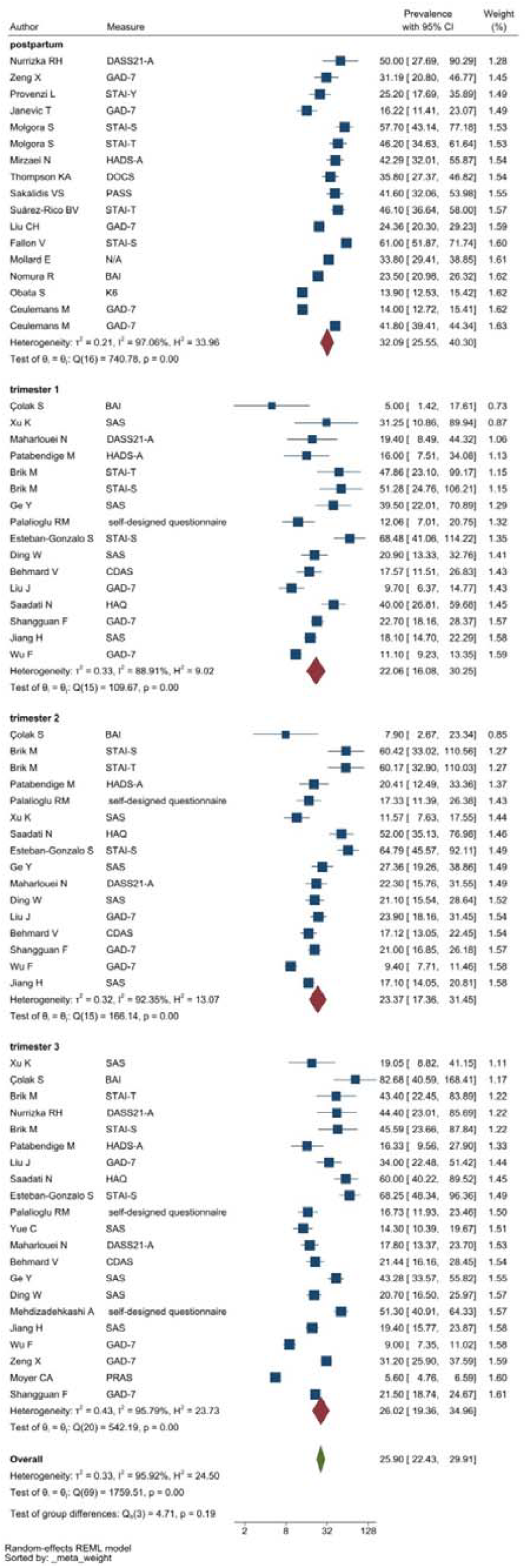
Subgroup analysis of anxiety (supplementary material)

**Figure 9.**
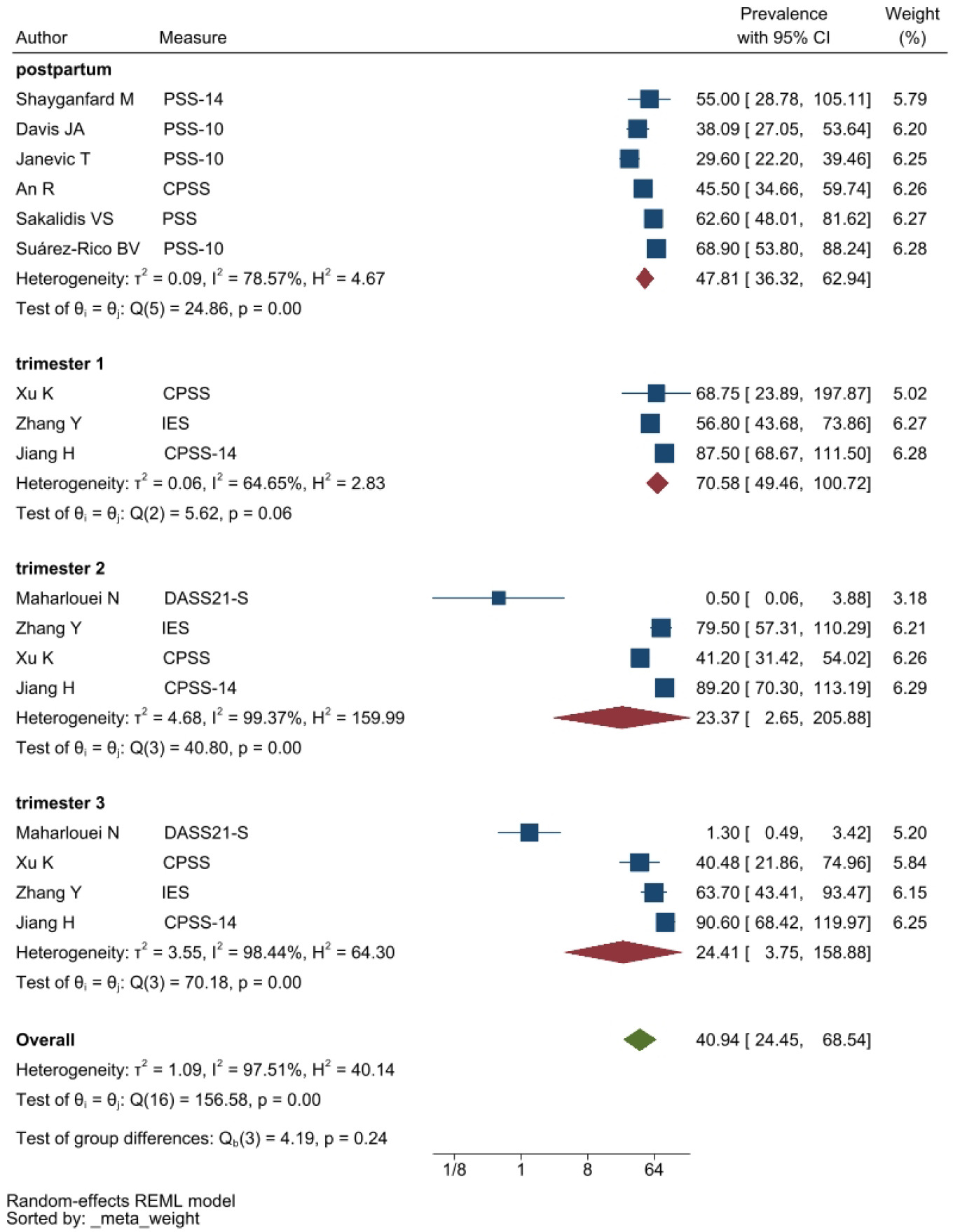
Subgroup analysis of stress (supplementary material)

**Figure 10.**
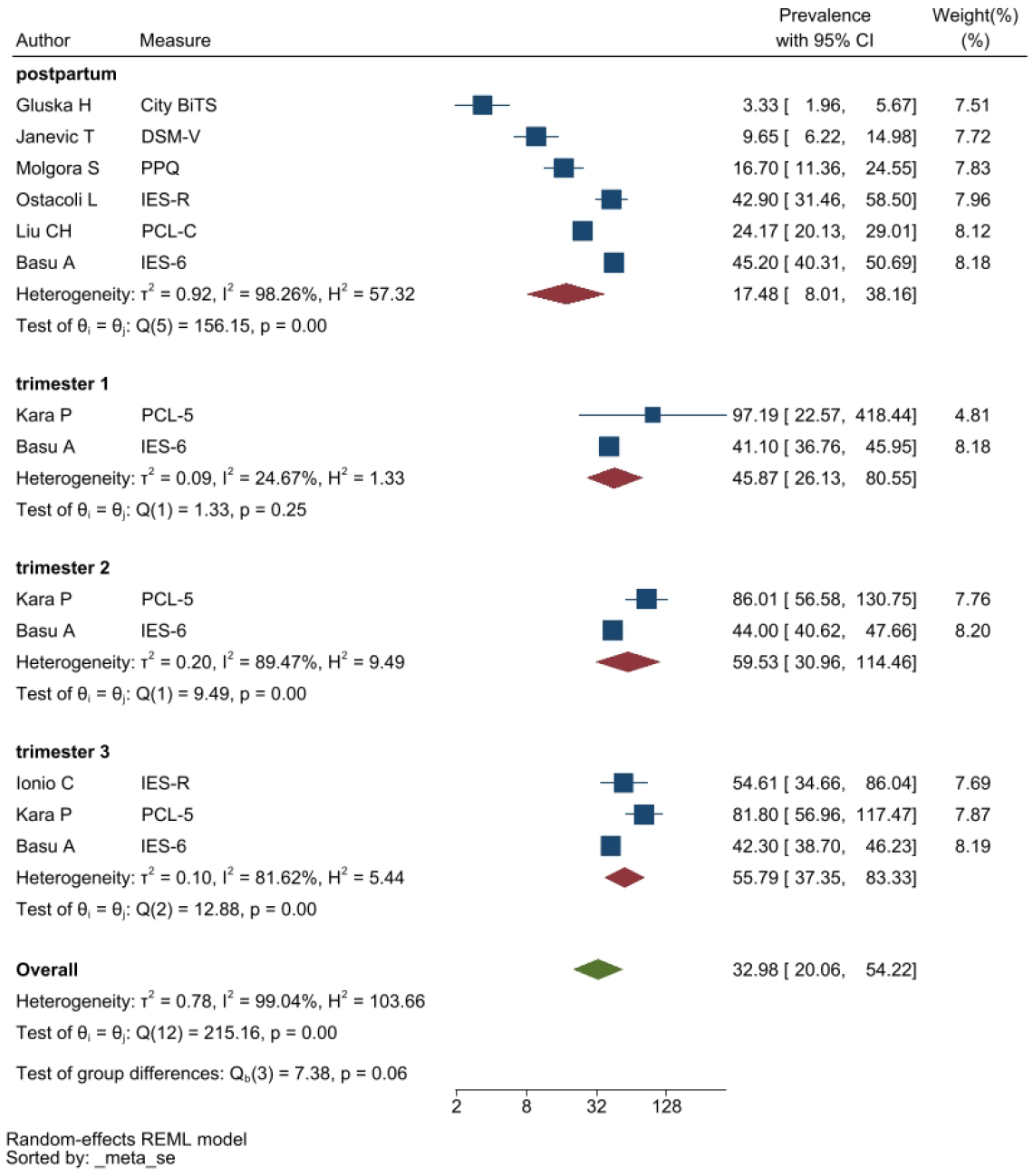
Subgroup analysis of PTSD (supplementary material)

**Figure 11.**
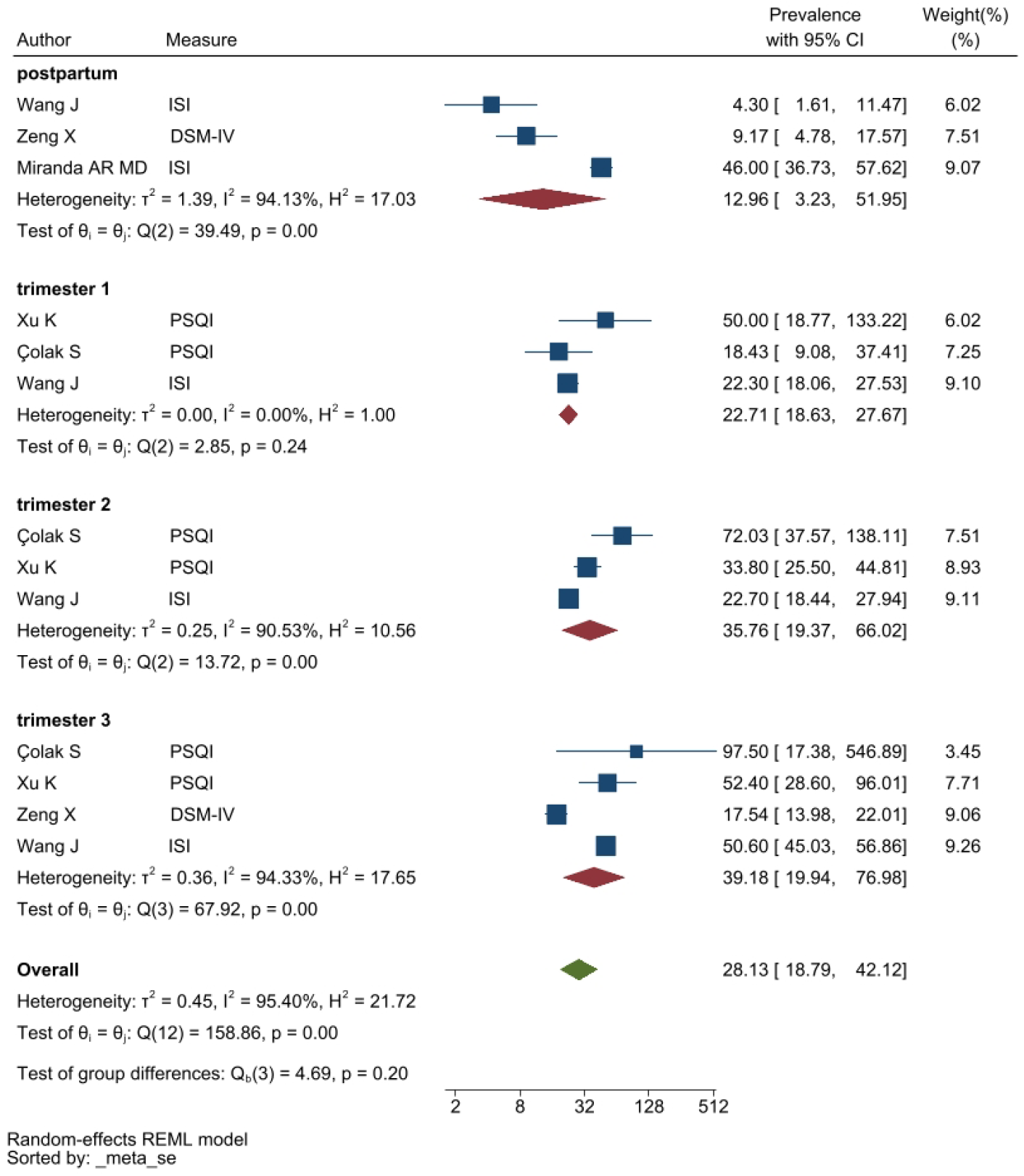
Subgroup analysis of sleep disorders (supplementary material)

**Figure 12.**
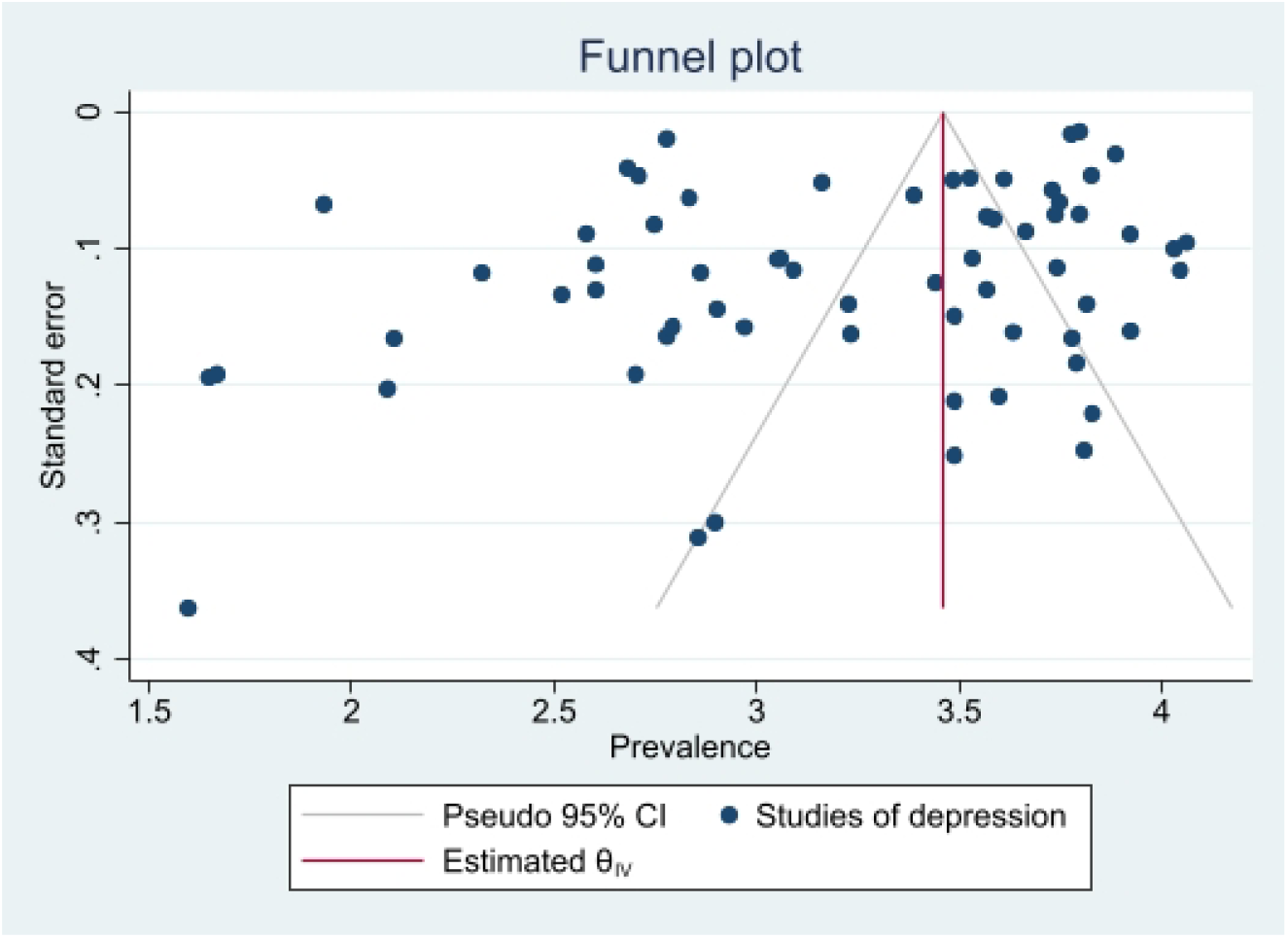
Funnel plot of depression.

**Figure 13.**
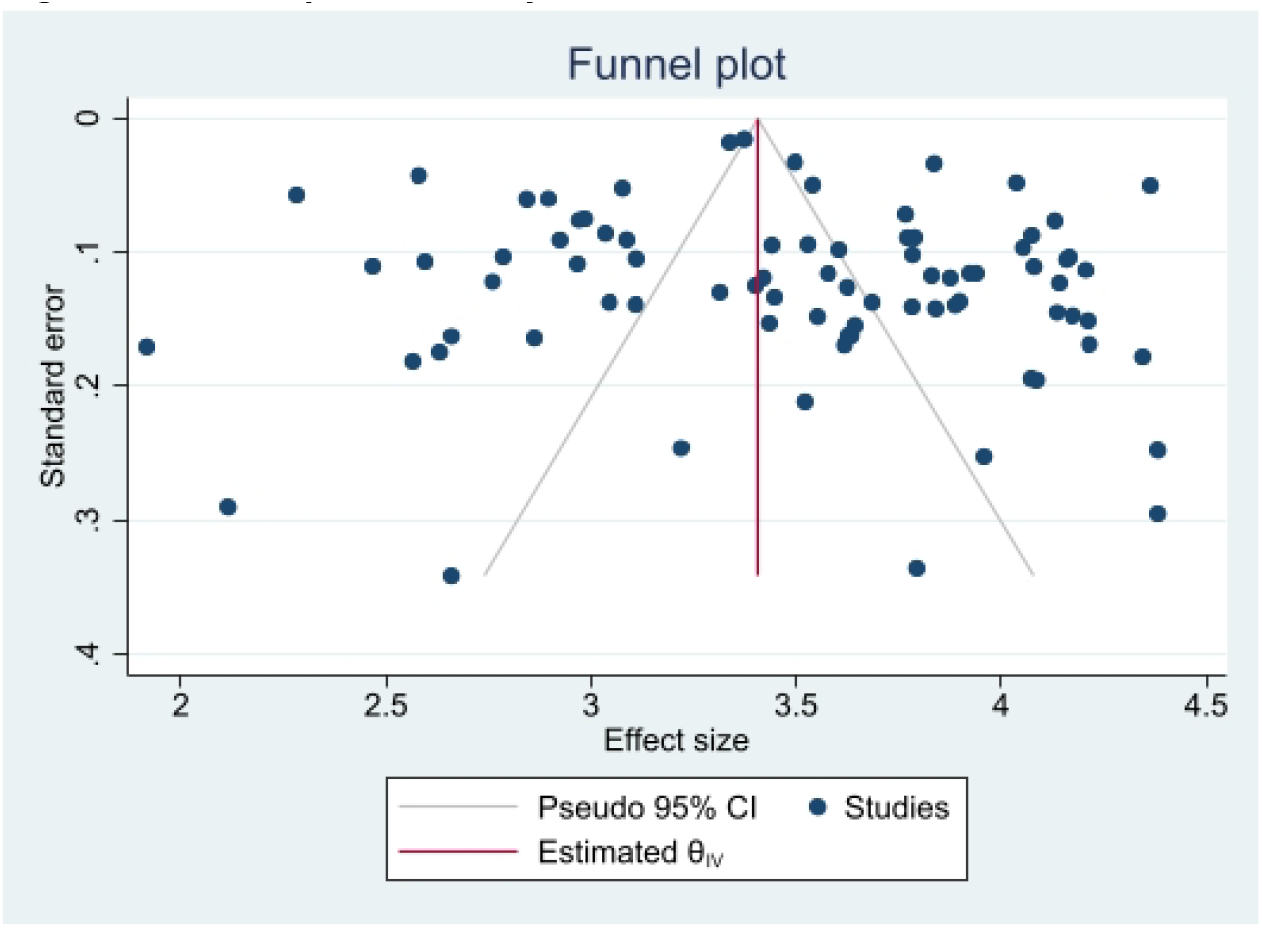
Funnel plot of anxiety.

**Figure 14.**
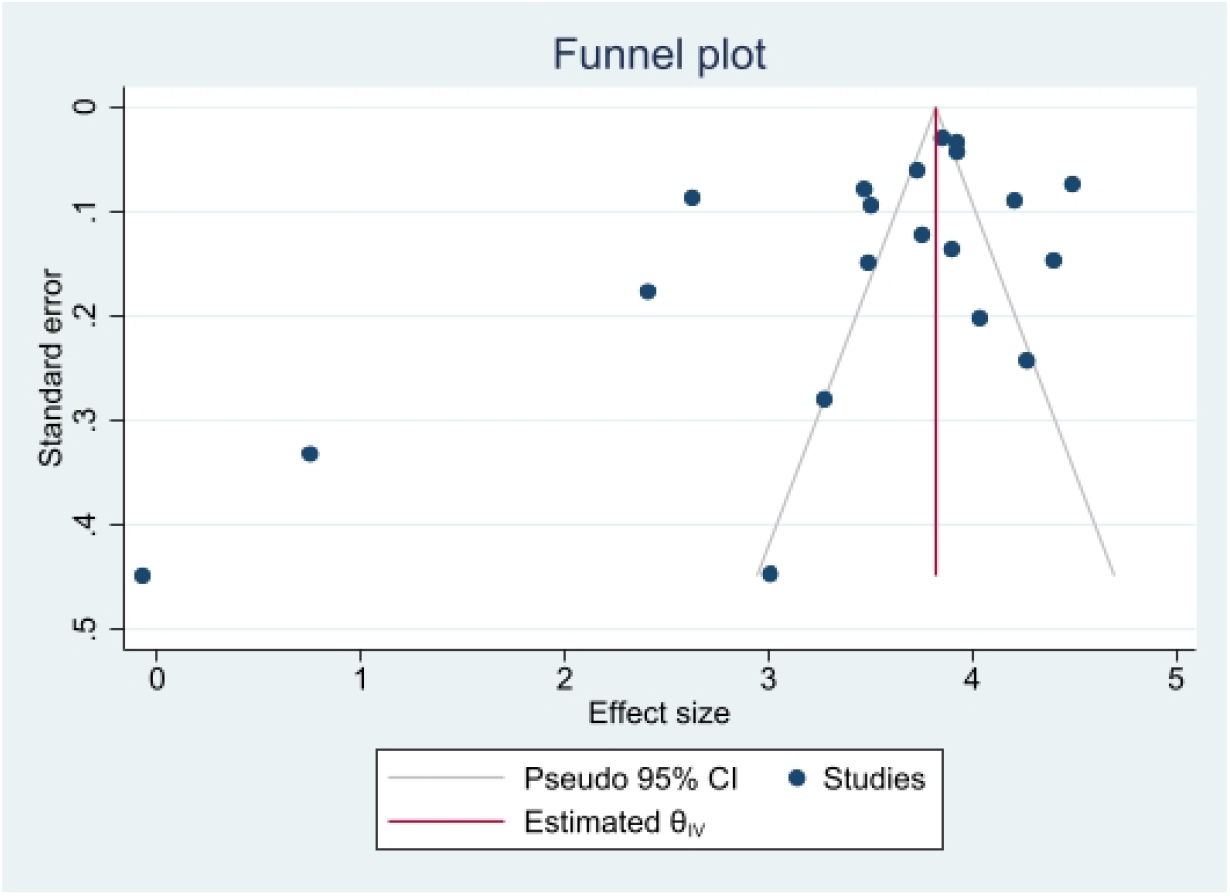
Funnel plot of stress.

**Figure 15.**
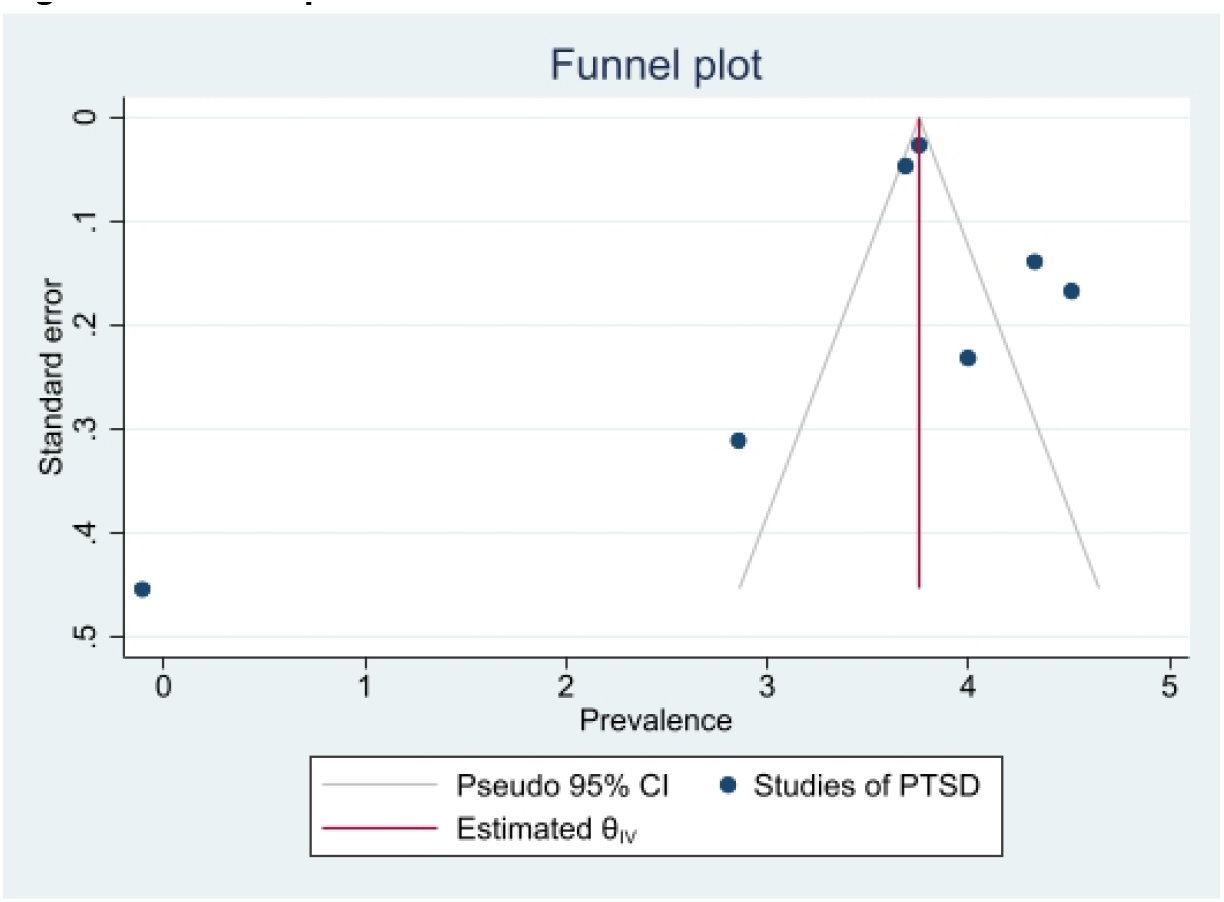
Funnel plot of PTSD.

**Figure 16.**
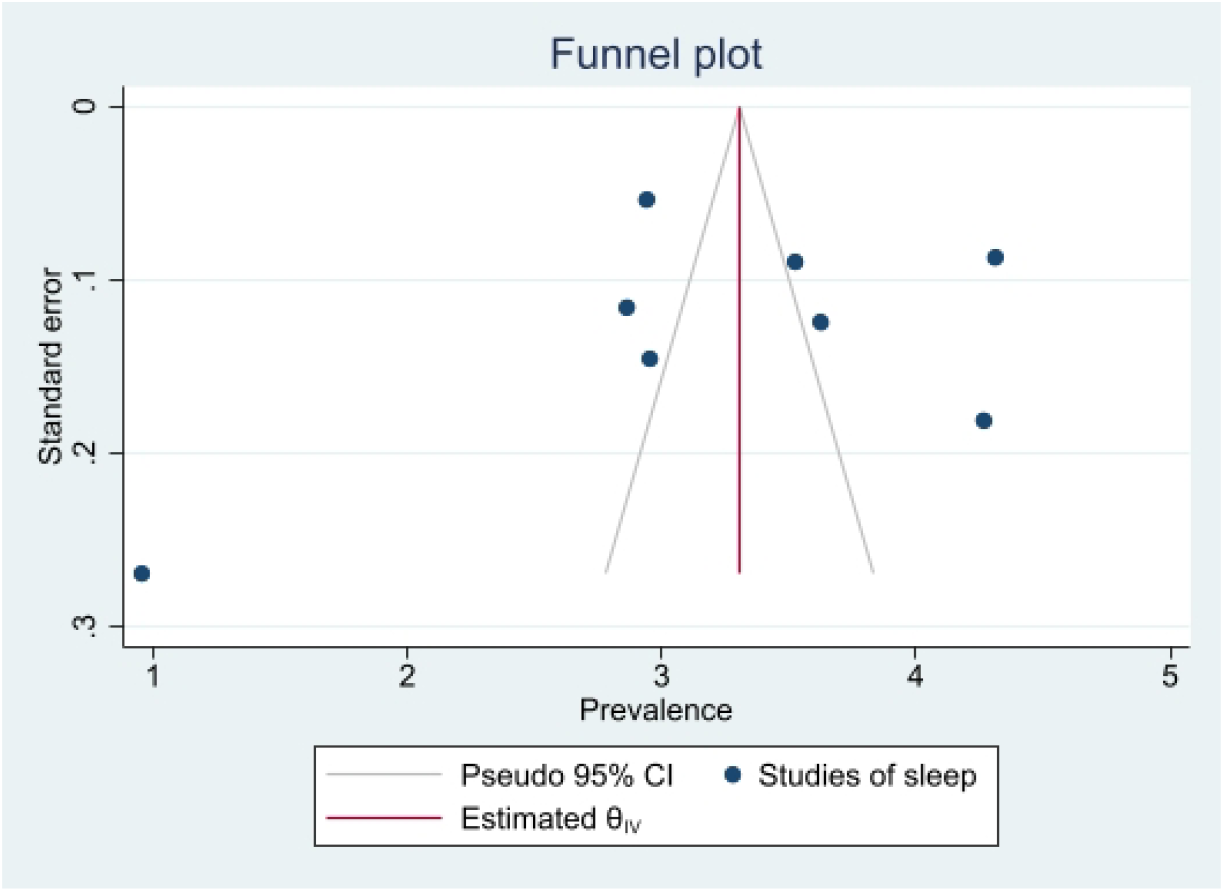
Funnel plot of sleep disorders (supplementary material)

**Figure 17.**
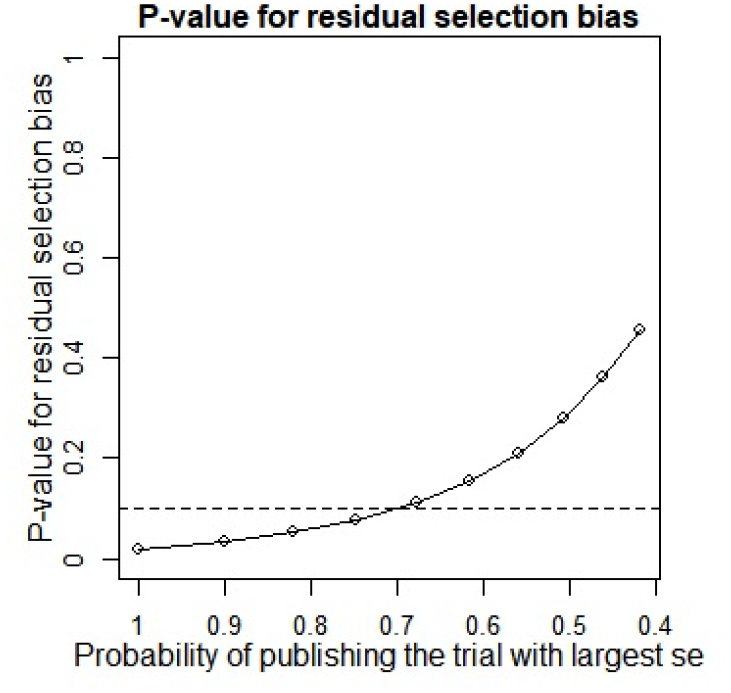
P-value for residual selection bias of depression.

**Figure 18.**
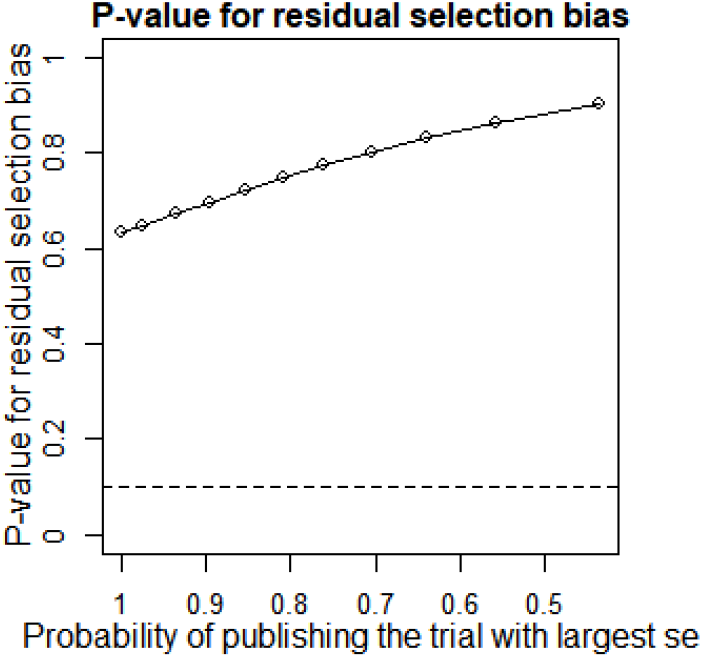
P-value for residual selection bias of anxiety.

**Figure 19.**
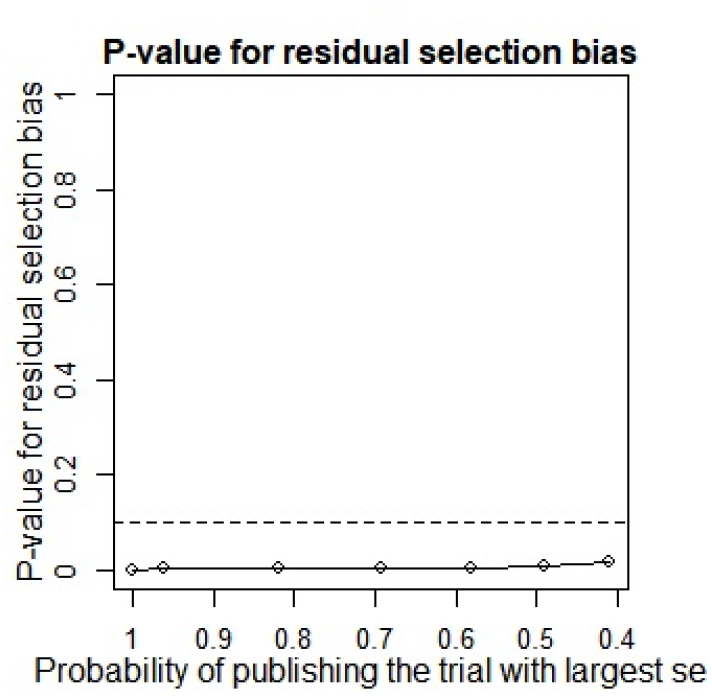
P-value for residual selection bias of stress.

**Figure 20.**
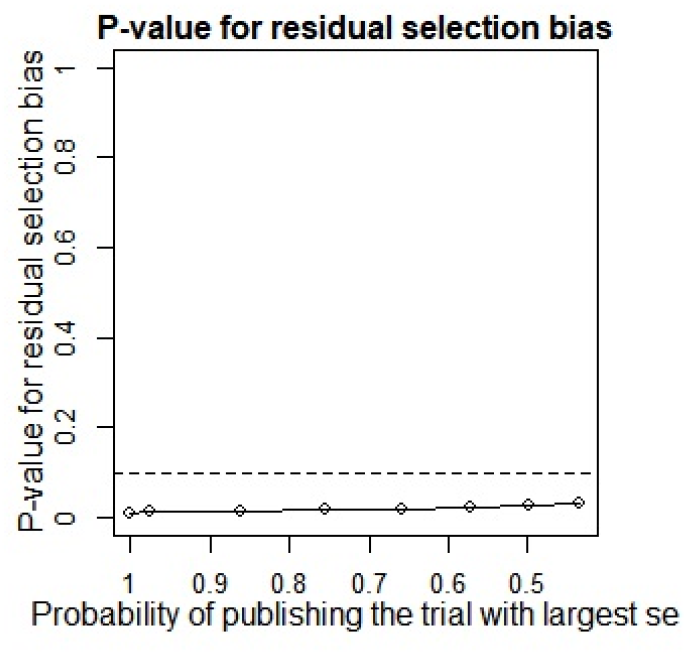
P-value for residual selection bias of PTSD.

**Figure 21.**
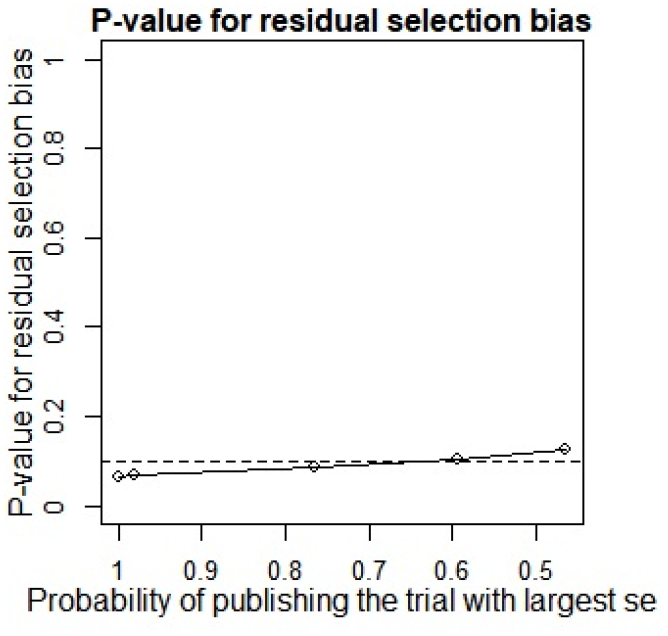
P-value for residual selection bias of sleep disorders (supplementary material)

A summary of studies used within the Copas selection model and random effects model has been demonstrated in Table 3, which indicates that the two models have no significant difference. P-value of the changes between these conclusions are 0.1108, 0.638 and 0.1042 for depression, anxiety and sleep disorders, respectively. The p-value of the Egger’s test was 0.0256 (**Table 4. supplementary material**) for studies of depression, revealing the existence of publication bias. The p-values of 0.256 and 0.998 (**Table 4. supplementary material**) indicates that it is challenging to detect publication bias for studies associated with anxiety and sleep disturbances.

**Table 3.** Summary of sensitivity analysis (supplementary material)

| Outcome | N of study | Model | Probability of publishing study with largest standard error | Proportion(%) | lower(%) | upper(%) | p-value for differences between two conclusions |
| --- | --- | --- | --- | --- | --- | --- | --- |
| Depression | 64 | copas selection model | 67.84% | 27.11 | 24.32 | 30.22 | 0.1108 |
|  |  | random effects model |  | 24.91 | 21.37 | 29.02 |  |
| anxiety | 82 | copas selection model | 100.00% | 32.88 | 29.08 | 37.18 | 0.638 |
|  |  | random effects model |  | 32.88 | 29.05 | 37.21 |  |
| Sleep disorders | 8 | copas selection model | 59.49% | 27.11 | 14.94 | 49.21 | 0.1042 |
|  |  | random effects model |  | 24.38 | 11.89 | 49.95 |  |

**Table 4.** P-value of Egger Test for the five mental health symptoms (supplementary material)

| Outcome | N of studies | p-value of Egger test |
| --- | --- | --- |
| depression | 64 | **0.0256** |
| anxiety | 82 | 0.256 |
| stress | 20 | 0.069 |
| PTSD | 7 | 0.742 |
| sleep disorders | 8 | 0.998 |

Note: ( \* ) : p<0.05 indicates significance

## Discussion

### Main findings

Our study demonstrates that depression, anxiety, PTSD, stress and sleep problems were common throughout the pregnancy period and after childbirth during the COVID-19 pandemic with 24.9% of women reporting symptoms of depression, 32.8% anxiety, 29.44% stress, 27.93% PTSD, and 24.38% sleep disorders. The lack of research conducted to assess the mental health impact of SARS and MERS on pregnant women is a significant limitation as such data could have supported preparation for similar pandemics in the future. Our meta-analyses indicated a clear-cut mental health impact of COVID on pregnant and post-partum mothers with a pooled prevalence of multiple symptomatologies of depression, anxiety, PTSD, stress and insomnia.

### Strengths and weakness

To our knowledge, this is the first systematic review and meta-analysis to focus on mental health outcomes in women during pregnancy and after childbirth during the Covid-19 pandemic. The searches were not limited by geographical location or language, therefore, further increasing the chances for all relevant literature to be identified. The MESH terms used did not consider all types of obstetric or gynaecology conditions but did include the common conditions. The variety of screening tools used across the included studies must be considered when interpreting the results of this review. Direct comparisons cannot be made where the same screening tool was not used. Furthermore, most studies used self-reported questionnaires, with no clinical follow-up to confirm diagnoses. Therefore, the results cannot be interpreted as prevalence of mental illness, but rather prevalence of symptomatology.

### Interpretation

Some studies have demonstrated that the extent and severity of mental health impact increased in women during pregnancy and after childbirth during humanitarian disasters and pandemics which is similar to our study [11].

The subgroup analysis showed that the prevalence of depression is identical during the first trimester of pregnancy [24.61% (95% CI 17.12 – 35.37)] and after childbirth [24.96 (95% CI 20.26 – 30.76)] compared to the second and third trimesters when the prevalence of depression is much lower at 16.52 (95% CI 9.31 – 29.33), and 22.49 (95% CI 18.91 – 26.74), respectively. This is suggestive of women who became pregnant and gave birth during the pandemic suffered from depression more frequently in the early stage and after birth, which appears to have been plateaued during the latter part of their pregnancy. It is unclear as to the reason for this observation, and the impact of this in a real-time scenario. The prevalence of anxiety, on the other hand, is higher among women after childbirth [32.09 (95% CI 25.55 – 40.30)], compared to an identical prevalence of anxiety during all the three trimesters of pregnancy [1^st^ trimester 22.06 (95% CI 16.08 – 30.25), 2^nd^ trimester 23.37 (95% CI 17.36 – 31.45), 3^rd^ trimester 26.02 (95% CI 19.36 – 34.96)]. This finding suggests that women after childbirth suffered more from anxiety during the Covid-19 pandemic. The stress level was significantly higher in women during the 1^st^ trimester of pregnancy 70.58% (95% CI 49.46 – 100.72), compared to 47.81% (95% CI 36.32 – 62.94).

This could be due to some of these women being first-time mothers or, general stress and health anxiety regarding how and when to access care from midwives and obstetricians as part of routine and emergency maternity care due to the Covid-19 pandemic. The findings of high level of stress amongst pregnant women is in keeping with other studies carried out during the Covid-19 pandemic that reported up to 70% of pregnant women suffered from stress during the pandemic[8]. Being pregnant and giving birth are known triggers for women to develop anxiety, and depression and pregnancy is a known risk factor for exacerbations or decline in pre-existing mental ill-health[9,10]. Other possible reasons for the increase in mental ill-health in women during pregnancy or after childbirth may be because of the massive clinical changes that took place regarding how women could access maternity care during the Covid-19 pandemic. As pregnant women were at higher risk of severe illness if they become infected with severe acute respiratory syndrome (SARS)-CoV-2 and develop COVID-19, pregnant women were advised to be stringent with public health measures such as social distancing and self-isolation to lower their risk of COVID-19 exposure. This led to the rapid implementation of virtual access to antenatal care to minimising the need for travel to antenatal clinics and in-person contact with healthcare staff, and antenatal care changed immediately from face-to-face consultations to telephone or video consultations. Birth partners were limited in number and visiting hours for partners were restricted resulting in less emotional and psychological support for women during labour in the delivery room, and after childbirth on the postnatal wards. Furthermore, as the Covid-19 vaccination was developed and the implementation programme initiated, there was uncertainty regarding the effectiveness and safety of the Covid-19 vaccine in women who were pregnant, which may have contributed and exacerbated stress and anxiety.

### Recommendations

All women should be risk assessed for maternal mental health at their booking visit and screened at every contact during pregnancy and after childbirth. All healthcare systems need to invest in perinatal mental health services delivered from a multi-disciplinary team including mental health nurses, specialist midwives, obstetricians with specialist interest in mental health and perinatal psychologists and psychiatrist. Maternity mental health services should be delivered in a way that meets the specific needs of the individual patient, including face-to-face consultations, telephone calls and/or video consultations. Up to date information regarding the impact of Covid-19 on maternity services needs to be available and easily accessible for women during pregnancy and after childbirth, for example by using social media campaigns and hospital websites. Learning from this data derived from COVID pandemic and consideration of the special needs of the pregnant and postnatal mothers should be imperative in strategies to implement early to improve preparedness of the health service in future pandemics.

## Conclusion

This study highlights that maternity mental ill-health was common during the Covid-19 pandemic and highlights the need to understand the complexity of factors associated with maternal mental health. Maternity mental health services need further investment and prioritisation and clear effective referral pathways and support for women who report mental health concerns during and after pregnancy are needed and require further research as to how best provide this care in a way that meets the specific needs of each women, across different healthcare systems.

## Data Availability

All data produced in the present work are contained in the manuscript

## List of abbreviations

MERS: Middle Eastern Respiratory Syndrome
SARS: Severe acute respiratory syndrome
EPDS: Edinburgh Postnatal Depression Scale
SAS: Self-rating Anxiety Scale
IES: Impact of Events Scale
ISI: Insomnia Severity Index
PSQI: Pittsburgh Sleep Quality Index
IAPT: Improving Access to Psychological Therapy

## Declarations

### Ethics approval and consent to participate

Not applicable for this review?

### Consent for publication

Not applicable?

### Availability of data and materials

Data availability statement goes here.

### Authors’ contributions

Authors’ individual contributions should be mentioned here.

## Acknowledgements

Authors should obtain permission from everyone to be acknowledged in this section.

